# Prioritising Research through Engagement with older Adult Care Homes and potential for mapping to existing data sources: the PREACH study

**DOI:** 10.64898/2026.10.01.26364274

**Authors:** Lisa Irvine, Kerry Micklewright, Bethany Myles, Megan Davies, Anne Killett, Claire Goodman, Jennifer Kirsty Burton

## Abstract

**Background:** Care home research often fails to address issues that matter most to residents, families, and staff. Priority-setting exercises can help reduce this lack of inclusivity, but few have received significant resident responses.

**Objectives:** To identify research priorities of UK care home stakeholders (including residents; relatives; care staff, managers and owners; healthcare professionals; policymakers and researchers), and to assess whether these priorities can be addressed using existing research datasets.

**Methods:** PREACH was an 18-month research priority-setting study involving 1063 stakeholders, across 50 care homes, including 381 residents. Participants submitted ideas of what care home research should focus on through 1) care home activity sessions or 2) an online survey. Responses underwent content analysis to identify themes, and these themes were mapped against a national repository of care home trial data.

**Results:** Fifty-one themes were identified. The most frequent priority across all stakeholder groups was access to meaningful activities for residents (22%). Relatives and residents emphasised social connection and day-to-day experience, whereas staff more often highlighted system pressures. No stakeholder-prioritised topics were directly addressable using existing secondary datasets.

**Conclusions:** Collaboration with care home teams to reach large numbers of residents offers a practical route to producing more relevant and timely care home research. Care home stakeholders consistently prioritise quality of life, meaningful activity and staffing over disease-specific or health-focused research. Existing research datasets measure relatively few of the priorities identified by residents, highlighting the need for resident-centred outcome measures and data collection.

## Background

Care home research can contribute to improving the lives of older adults living there, and the staff who care for them [1]. To ensure valuable resources are not wasted and research is relevant [2], researchers must address the uncertainties and questions that matter most to people living and working in care homes. Systematically capturing these perspectives in the form of a research priority setting exercise can generate useful evidence for researchers and funders.

Priority setting exercises often produce findings that require substantial further work before they can be translated into answerable research questions [3], and much of the existing literature focuses on engagement methods rather than how priorities can be operationalised [4]. Mapping stakeholder-identified priorities onto existing data sources can maximise the value of completed studies, and help ensure new analyses address areas of genuine importance to older adult care home residents, staff, families and other key stakeholders [5]. This is the central methodological innovation of the *Prioritising Research through Engagement with older Adult Care Homes* (PREACH) study [6].

The overarching aims of the PREACH study were to: identify important research priorities in older adult care homes; and determine if existing data could be repurposed to address the identified issues.

## Methods

The PREACH study collected research priorities through: an online survey, accessible to anyone with an interest in care home research; and in-person activities conducted with residents, designed to be facilitated by care home activity staff.

### Online survey

A brief online survey (JISC Online Surveys platform, see Appendix A1) was developed, broadly following methodology commonly applied in the UK [7]. Participants were provided information about the study, asked to provide informed consent, and self-categorised as: care home resident; care home staff member; family or friend of care home resident; health care worker; care home owner or manager; social services worker; researcher; policymaker or other. The range of priorities was intentionally broad, and no particular research design, topic or type of research question was specified. To collect the ideas, there were three free-text boxes with instructions *“Please add your ideas for future research in care homes”*. A promotional guide and materials including downloadable posters and leaflets were available online [8] (Appendix A2).

### Care home activities

In this paper, the term ‘care home’ is used as an overall term for residential environments providing permanent, 24-hour care to older adults [9]. To involve people living in care homes, the research team collaborated with two care home activity co-ordinators to co-design activity packs that could be used to enable discussion of research priorities; activity co-ordinators trialled these packs with small groups of residents, whose feedback was incorporated into subsequent amendments [10] [11]. Posters and leaflets (printed and digital formats, Appendix A3) were produced to promote study participation; designs were informed by previous Patient & Public Involvement and Engagement (PPIE) feedback [12].

Activity packs, promotional materials and prepaid envelopes were posted to interested care homes following consent from the manager. Resident engagement sessions were delivered by care home staff or NIHR-funded staff, using activity packs and suggestion cards. Postcard-sized cards were provided for residents to complete during group discussions and also delivered with information leaflets to each resident’s bedroom, enabling participation of residents who opt-out of group activities. The cards stated, *“My idea for care home research is…”* and asked respondents which of the listed stakeholder groups they belonged to. We assumed each returned card represented one individual contribution. To incentivise the return of responses, care homes could enter a prize draw. All care homes were offered certificates to recognise their participation.

Care homes were recruited via the NIHR Research Delivery Network (RDN), which facilitates research by offering resources, recruitment assistance, data collection, and advice. A small number of homes completed the activity without RDN support. Twice-weekly online drop-in sessions were scheduled for care home staff and RDN teams to seek advice. The research team also attended events organised by specific care homes including relatives’ meetings, and local RDN meetings.

## Analysis

### Content analysis

Survey and postal responses were combined. Qualitative content analysis was used to systematically code individual contributions and generate themes [13]. We employed a combined deductive and inductive coding framework, initially using themes derived from Graham et al. (2024) [14] and expanding the framework as new stakeholder priorities emerged. The coding process was iterative, involving regular team discussions to resolve uncertainties and maintain an analysis log for transparency. Unlike consensus-based models, prioritisation was determined by the frequency of mentions, with the most common topics regarded as having the highest shared importance. The team transitioned from NVivo (coding) to Microsoft Excel (analysis) to facilitate easier task-splitting and data sharing given the high volume of brief textual responses. Because the study focused on generating stakeholder-led ideas without constraints, no second-stage prioritisation exercise was conducted. More detail is provided in Appendix A4.

### Mapping to existing data sources

Contributions were initially classified according to whether they represented an original research idea or a general comment, concern or observation. An idea was considered a ‘*viable research question*’ when it went beyond describing a concern and identified an issue that could, in principle, be investigated through systematic research. Ideas that could not be reasonably expressed as a researchable question were classified as ‘not viable research questions’ and retained in the overall dataset but excluded from the subsequent assessment of potential secondary analysis. To assess whether stakeholder-identified priorities could be examined using existing secondary datasets, each *‘viable’* research question was reviewed to determine its conceptual alignment with variables available within the *Virtual International Care Homes Trials Archive* (VICHTA), a repository of individual participant data (∼6000 residents) from UK care home trials [15, 16].

Feasibility was assessed independently by two researchers with detailed knowledge of VICHTA (XX XX), with disagreements resolved through discussion with the wider team. This structured approach aimed to identify questions suitable for secondary analysis (rather than to reprioritise the questions according to data availability), highlight areas requiring methodological development, and inform future data collection priorities in care home research. More detail provided in Appendix A5.

### Ethical considerations

Ethical approval was provided by the University of XXX Health, Science, Engineering and Technology Ethics Committee with Delegated Authority (HSK/SFXXX). Respondents were considered public involvement contributors to the priority setting exercise, rather than research participants: no personally identifiable information was required or recorded, and any potentially identifying information in free-text responses was removed. Care home staff were asked to only involve residents who they judged to have capacity to engage meaningfully with the activities.

We report our findings in accordance with REPRISE [17], Appendix A6.

The entire dataset of deidentified stakeholder responses is publicly available as an open-source resource, which can be downloaded from www.carehomepriorities.com.

## RESULTS

### Response rates

The survey was open from 01/07/2024-30/06/2025, across the UK. There were 410 responses, self-identifying as: care home staff (n=123); healthcare worker (n=70); owner or manager (n=62); family or friend (n=62); researcher (n=50); resident (n=28); policy & other (n=8); social services worker (n=7).

Care home activities ran between January and July 2025, RDN teams in eight regions of England invited care homes to participate. Ninety-nine expressed interest and managerial consent was sought. Seventy-one consent forms were returned, and fifty care homes took part. Suggestion cards were completed and returned by n=353 residents, n=223 staff, and n=77 family/friends (See Figure 1 & Appendix A7).

**FIGURE 1.**
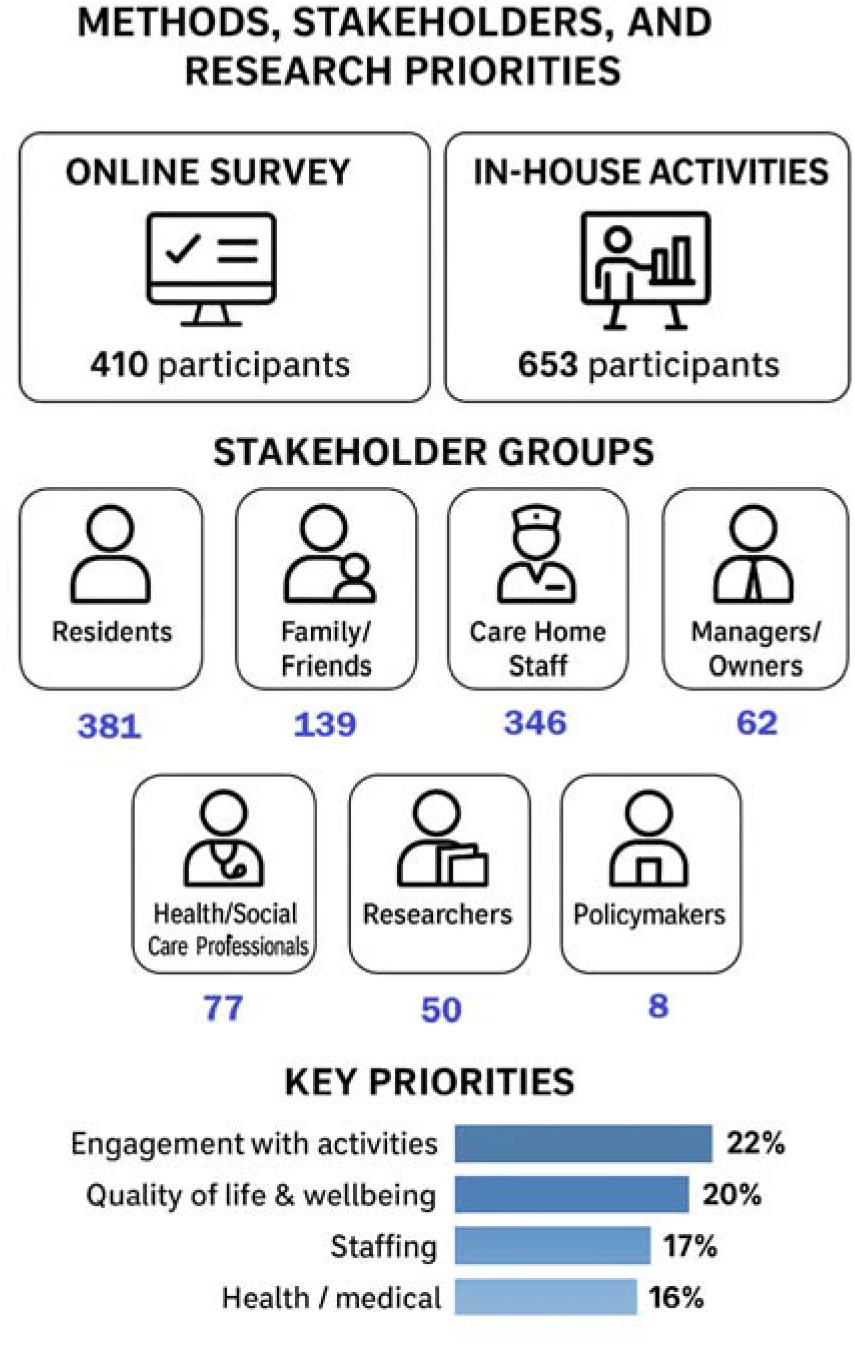
PREACH Framework

Due to the anonymous nature of the study, demographic makeup of respondents is unknown. The 50 care homes participating were largely representative, spanning private (n=39), voluntary (n=8), and local authority sectors (n=3) with mixed regulatory ratings. They varied in size, included both urban and rural locations, and 60% offered residential care only. Large chains were under-represented (6% sample:19% nationally), as were smaller homes (24% sample: 59% nationally) (Appendix A8). [18]

Updates were sent to care homes and RDN teams throughout the study, including a final “Headline Results” newsletter (Appendix A9).

### Identified priorities

Online survey and suggestion card data was combined, with 1063 total responses. Fifty-one descriptive themes were developed.

The most common themes across the data were “Engagement with activities”, “Quality of life and wellbeing”, “Staffing” and “Health or medical”. Here we summarise the top four major themes (See Figure 2, Appendices A10-11).

**FIGURE 2.**
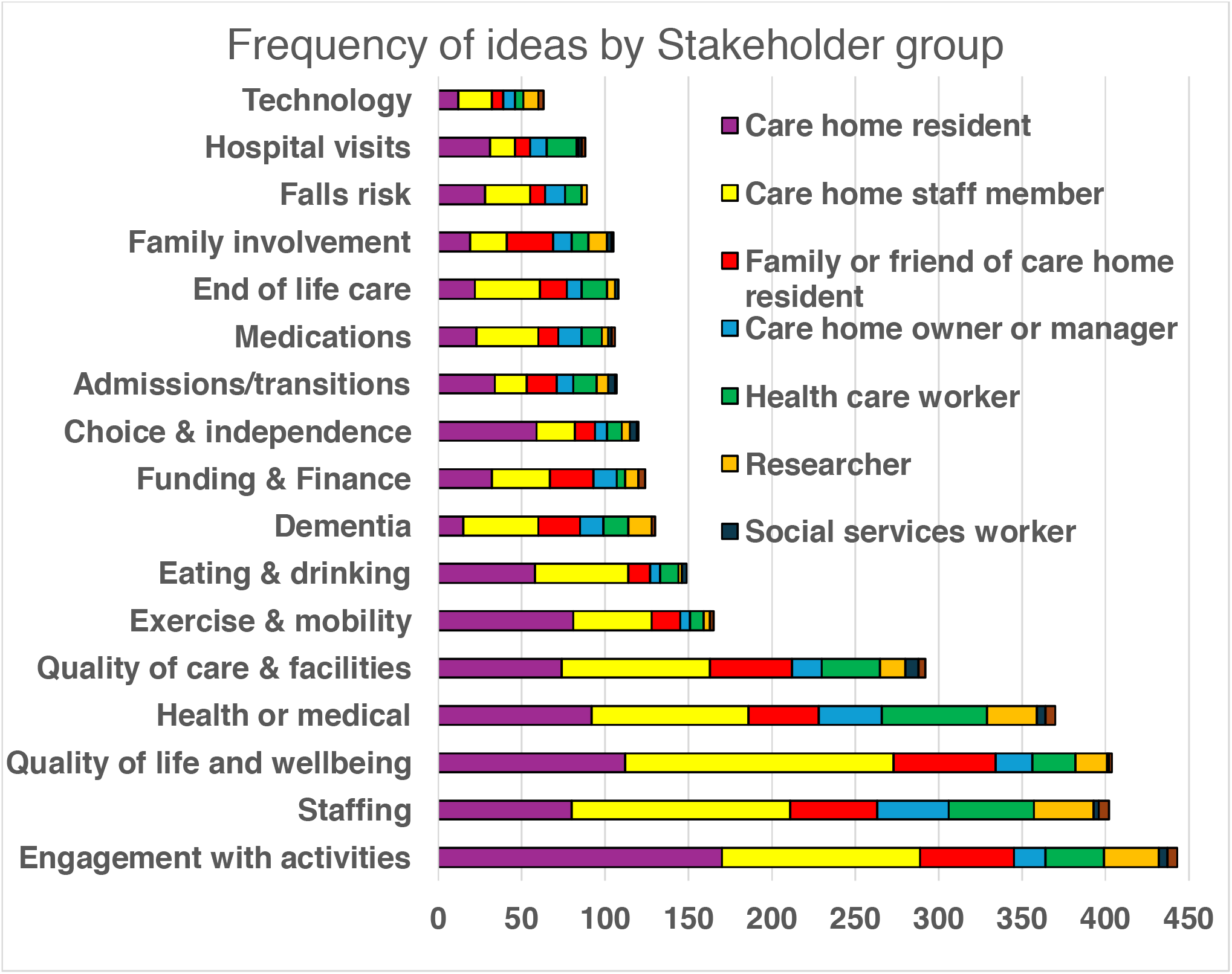
Frequency of idea (any mentions) by Stakeholder group

#### Meaningful Activities

Engagement with activities was the most mentioned topic overall, with 443 related comments (22% of the total suggestions). Seventy-seven unique activity ideas were discussed, broadly categorised as: Exercise; Social; Excursions; Cognitive/Learning; Creative; Inclusive and Other (See Table 1). Residents shared their desire to keep learning, including developing new skills and finding out more about medical conditions experienced within the home, such as dementia. Family members wanted residents to be able to get outside. Creating a sense of community and inclusivity was a common thread throughout the received responses.

**TABLE 1.**
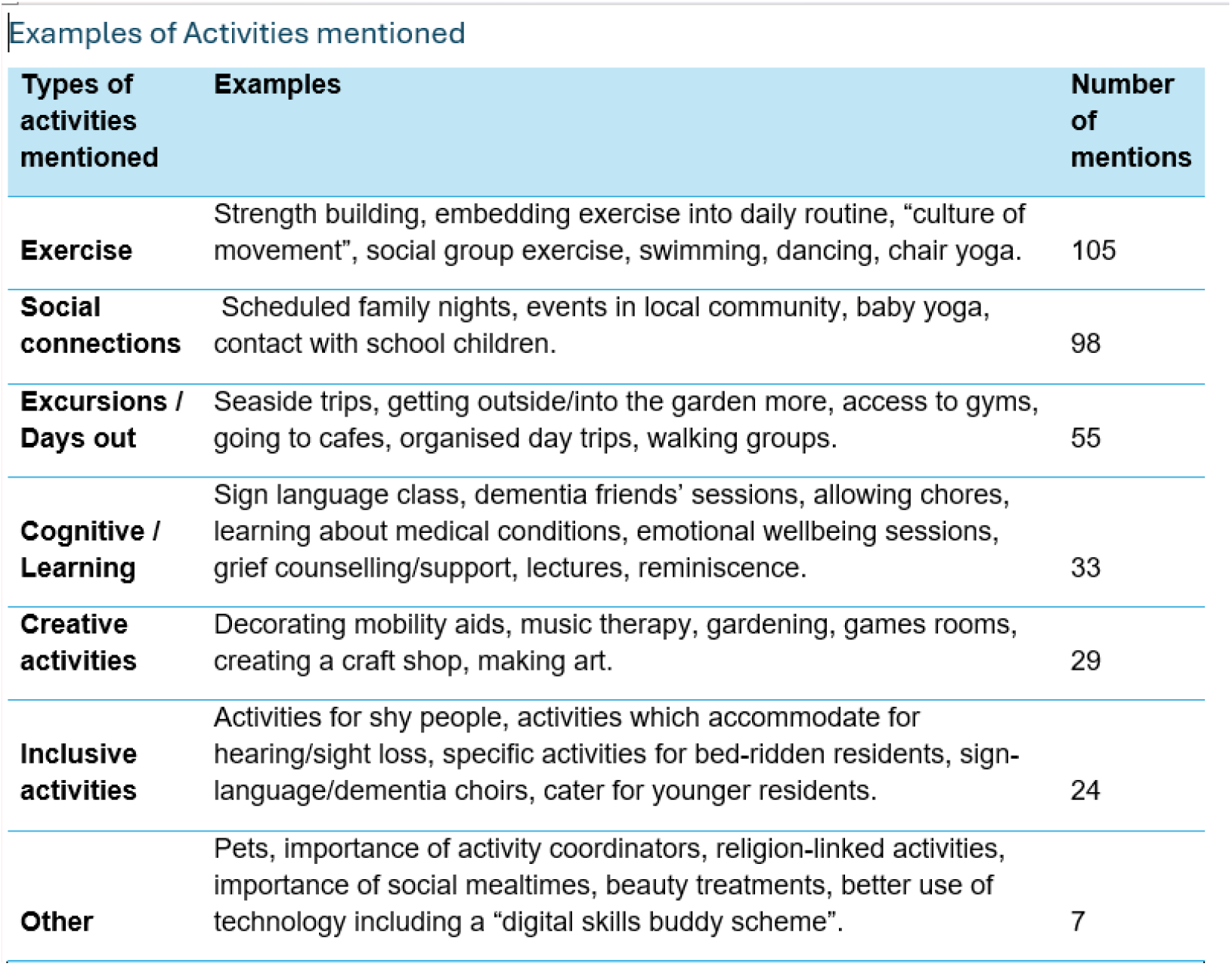
Types of Activities suggested.

**TABLE 2.**
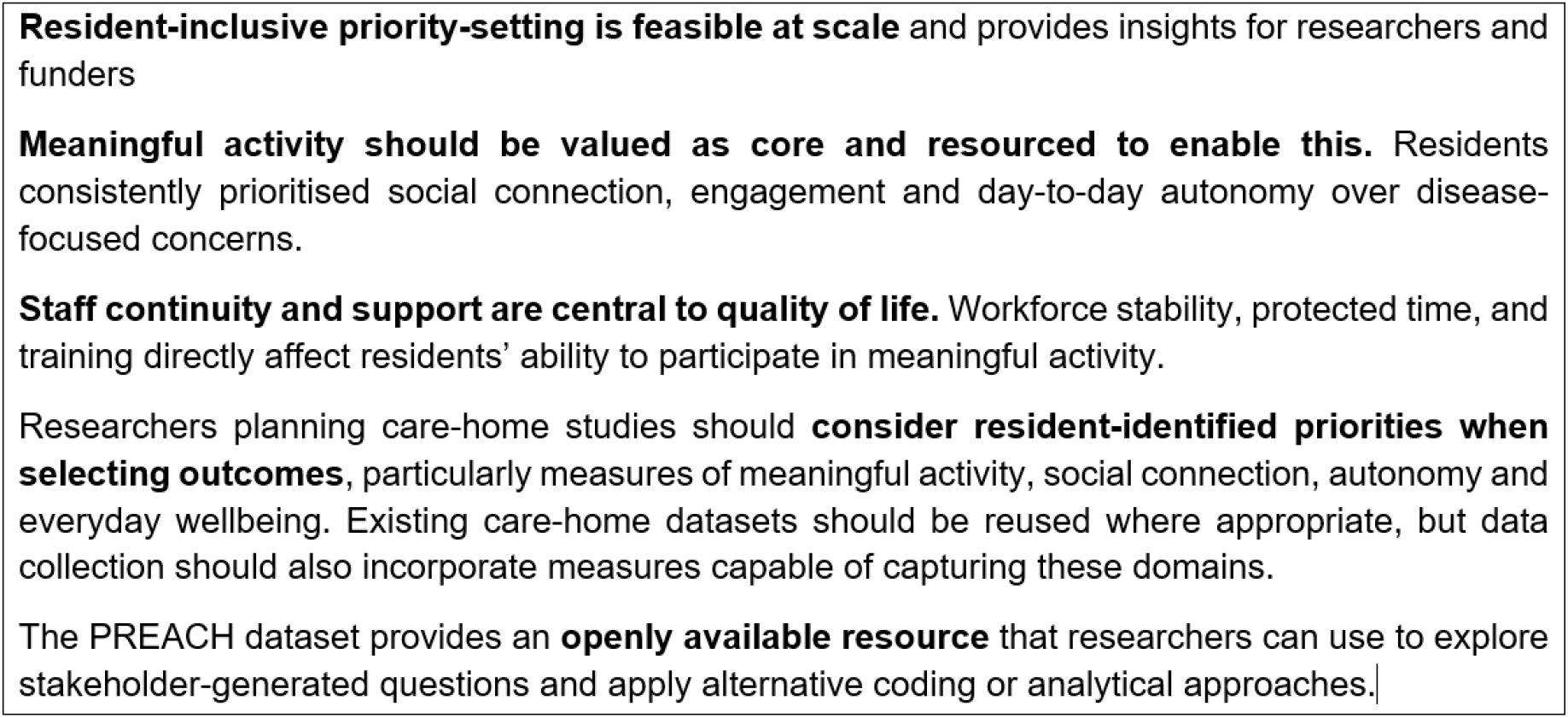
Implications for policy, practice and research.

#### Quality of Life / Well-being

Quality of life and wellbeing was discussed by 406 participants, particularly staff members and residents’ friends/relatives. Sub-themes related to resident choice, autonomy and independence, as well as specific discussion around social connection and loneliness. Residents wrote about having access to counselling and adjusting into care home living. Some care home residents wrote about not currently having access to age-appropriate activities as younger adults.

#### Staffing

Staffing was the most reported theme by owners or managers, policymakers, and researchers, with 402 responses overall. Care home staff raised concerns around staffing levels, pay levels, mental health and wellbeing, compassion from management and staff having sufficient training, while managers addressed staff retention and preparedness to work in the industry. Many responses from care home residents indicated awareness of recruitment challenges, and praised staff.

#### Health / Medical

Topics categorised in the “Health / medical” theme included difficulties accessing services, challenges relating to hospital discharge, and the burden of organising transport for appointments. Specific health concerns included wound healing, dysphagia, managing diabetes, supporting residents with Parkinson’s and maintaining continence.

Managers/owners were concerned with the prevalence of polypharmacy and reluctance to stop medications, even when they may no longer be needed. Care home staff wanted more planned, on-site clinical input (especially from GPs). Concerns were raised about deconditioning in care homes and potential discrimination against residents by NHS services.

### Comparing stakeholder groups

Our substantial response rate provided the opportunity to consider both shared and contrasting prioritisation between stakeholder groups. Residents and families (n=520) highlighted human connection, wellbeing and activities, while staff (n=346) considered practical challenges like retention and training. Care home owner/manager (n=62) priorities often related to operational and financial pressures, staff management, and external service interfaces. Researchers (n=50) focused proportionately more on dementia, other health-related topics, and technology. Social services workers (n=7), a much smaller group, focused on the rights, dignity, and individuality of residents, critiquing systemic failures, and challenging institutional practices. (Appendix A12)

### Mapping viable research questions to existing data sources

Our 1063 respondents contributed 2013 individual entries. Of these, 1528 were classified as original research ideas, 479 were considered themes or general comments and six could not be classified. Of the 1528 research ideas, 294 (19%) met the predefined criteria for a viable research question. These comprised 245 questions requiring primarily qualitative or observational research, 25 ideas for randomised controlled trials and 24 questions judged to have potential for secondary analysis using existing data (See Figure 3 & Appendix A5/A13/A14). No priorities were judged to be fully addressable using VICHTA data alone. Priorities that could be partially explored through existing variables included areas such as falls, medication use/safety, physical function/mobility, and hospitalisation/emergency care use. Priorities requiring new primary data collection were predominantly resident- and family-generated priorities and were concentrated within domains relating to meaningful activity, social interaction, mealtime experience, staff continuity, and wellbeing.

**FIGURE 3.**
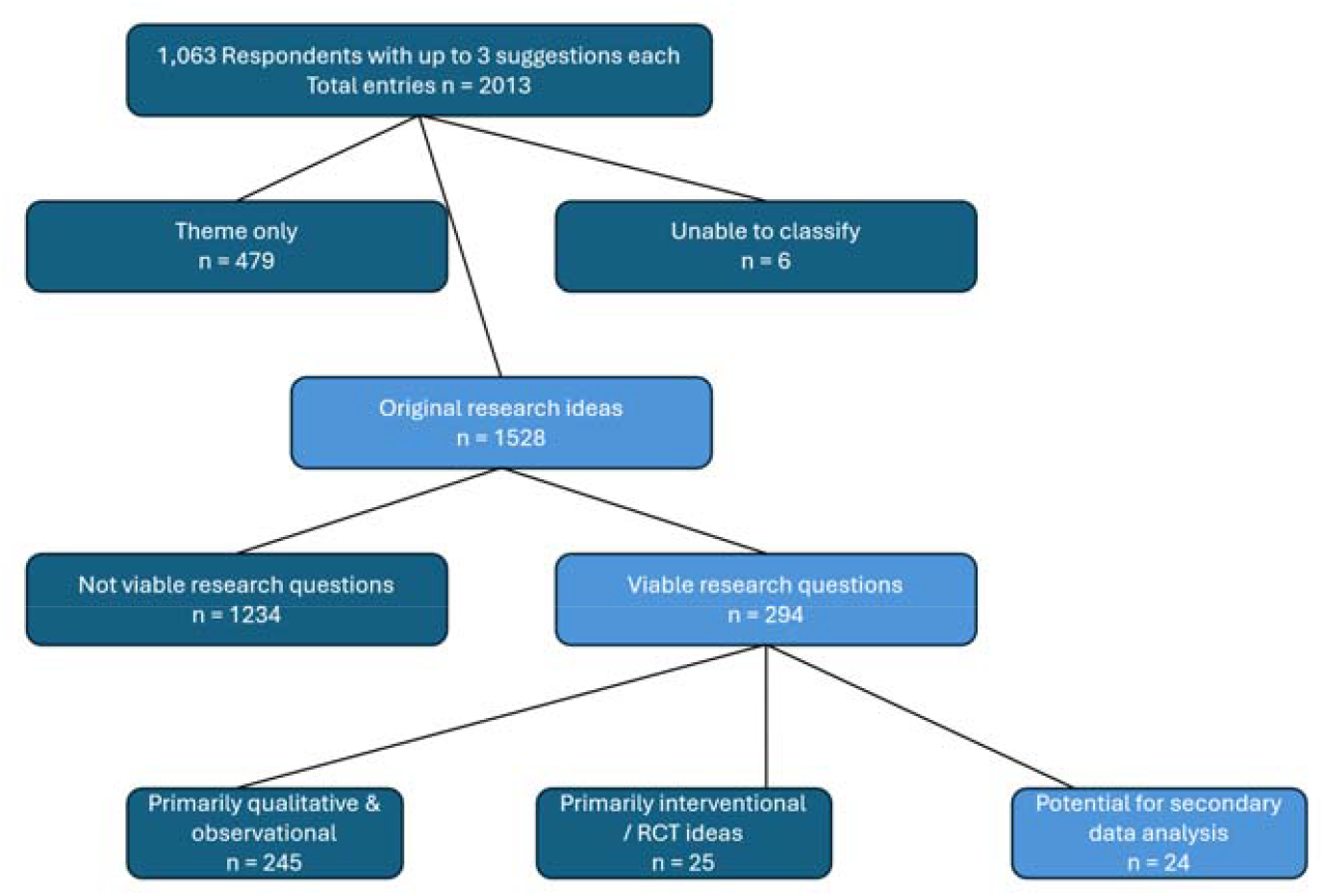
Flowchart of stakeholder suggestions & viable research questions

## Discussion

The PREACH study identified priorities shared by 1063 stakeholders, including a high proportion of care home residents. Our findings indicate that care home stakeholders placed strong emphasis on aspects of everyday life, while clinical concerns were less prominent. Residents’ responses focused on meaningful activities, social connection, autonomy and overall quality of life. All stakeholders noted persistent challenges with fragmented health services and staffing pressures. Recent systematic reviews [14, 19], show that most published priorities focus on clinical outcomes, health-related behaviours and health-system topics. PREACH placed particular emphasis on resident participation, generating priorities rooted in lived experience. The importance of meaningful activities for residents is strongly aligned with findings from a recent priority setting exercise in the UK [20], which also demonstrated the feasibility and value of engaging residents, their relatives, and frontline staff in shaping research agendas. Compared with Shepherd [21], where person-centred care dominated staff-generated priorities, this concept appeared far less frequently in our data. PREACH echoed Australian findings [22] on staffing and funding pressures, but provides a greater focus on what care home priorities look like when residents themselves are placed at the forefront.

A methodological innovation of the PREACH study was its systematic mapping of stakeholder-generated priorities to existing data sources, such as the VICHTA trials archive. This approach has the potential to reduce research waste by making fuller use of routinely collected health and social care data, and has proven highly effective in fields like stroke, where trials share standardised clinical and functional endpoints [23]. In practice, however, very few stakeholder-identified priorities could be addressed using existing UK care home trial datasets. Despite the availability of pooled individual participant data through VICHTA, a relatively small proportion of high-priority topics were directly or even partially addressable without new primary data collection. This highlights a mismatch between stakeholder priorities, and the outcomes represented in existing care home trial datasets [24], with implications for future research commissioning. Many resident priorities concerned aspects of everyday life that were not captured by the outcomes available in VICHTA. While pooled trial datasets such as VICHTA are well suited to analysing clinical outcomes and service use, they offer limited capacity to address the relational and experiential domains prioritised by residents. Limited alignment observed is not a failure of engagement or priority setting, but a consequence of what is routinely measured. Aligning research agendas with resident priorities will require parallel investment in developing and embedding resident-centred measures suitable for large-scale data collection [25].

The emphasis residents placed on activities was a striking finding. However, the activity-based format of data collection may have influenced how priorities were expressed, as participants’ responses can be shaped by the context in which they are elicited. Variation in how activities were delivered across homes meant that fidelity could not be formally assessed, and local facilitation may have shaped the framing of questions. Nevertheless, the prominence of activities in our findings aligns with previous work [20], which was not explicitly activity-led, suggesting that this emphasis reflects genuine resident concerns rather than methodological artefact.

When comparing priorities of different stakeholder groups, dementia was mentioned less frequently and in a different context by residents than by researchers and other professional groups Researchers gravitated toward topics that are “tangible”, methodologically tractable, and well-aligned with existing ring-fenced funding frameworks (e.g., dementia or digital technology).

### Strengths and limitations

A key strength of the PREACH study was the scale and breadth of participation, successfully engaging 1063 stakeholders across 50 care homes and securing responses directly from 381 older adult residents. By collaborating with frontline activity coordinators to co-design tactile, accessible engagement materials, the study demonstrates the feasibility of participatory research within long-term care settings [11]. Additionally, the systematic mapping of priorities against the VICHTA trials archive represents a distinctive methodological innovation to reduce research waste.

Several limitations must, however, be acknowledged. First, all submissions were completely anonymous. While this maximised participation and ensured GDPR compliance, it prevented the research team from seeking follow-up or clarification on ambiguous suggestions. No individual-level demographic data were collected, which limits our ability to formally assess equity or determine which specific groups were underrepresented.

Our methodology introduced specific participation and facilitation biases. To ensure ethical compliance, facilitators only invited residents assessed as having capacity to engage with the activities. Consequently, the views of residents with advanced dementia or severe cognitive impairment, who represent a significant portion of the long-term care population, are not captured. While bedroom-delivered cards were provided to enable independent, private feedback, these were rarely completed, meaning most resident responses were transcribed or facilitated by activity coordinators. This active facilitation, combined with conducting the sessions during scheduled group activities, may have primed residents to prioritise “meaningful activities”.

Prioritisation based on ‘frequency of mentions’ provided a transparent approach for summarising a large volume of stakeholder contributions, but should not be interpreted as equivalent to consensus or as indicating that less frequently mentioned issues were less important. In addition, translating stakeholder contributions into viable research questions required researcher interpretation, including decisions about whether contributions represented researchable questions and how related ideas were categorised. Finally, PREACH did not systematically determine whether identified priorities had already been answered by the wider evidence base. The findings should therefore be interpreted as a stakeholder-informed assessment of research priorities and their potential for secondary analysis, rather than a definitive ranking of research needs or evidence gaps.

## Conclusions

The PREACH study shows that large-scale, resident-inclusive engagement is feasible and informative, identifying research ideas that reflect life in care homes. Stakeholders identified many priorities concerning relational and experiential aspects of care home life, but existing trial datasets were poorly equipped to answer them, highlighting the need for new primary research and better measurement of outcomes reflecting residents’ everyday experiences. Without these changes, care home research risks continuing to prioritise what is most readily measurable and health specific, rather than what residents and staff identify as most important.

## Data Availability

The PREACH dataset is publicly available (EEPRU weblink) to support further research, and we encourage researchers to use it to explore their own areas of interest and maximise the value and impact of the data collected.

https://www.carehomepriorities.com/

## Data availability

The PREACH dataset is publicly available (XXX weblink) to support further research, and we encourage researchers to use it to explore their own areas of interest and maximise the value and impact of the data collected.

## References

1. National Institute for Health and Care Research, N. Advancing Care - Research with care homes. 2017 [cited 2025 15 December]; Available from: https://evidence.nihr.ac.uk/collection/advancing-care-research-with-care-homes/.

2. Peryer, G., Kelly, S., Blake, J., Burton, JK., Irvine, L., Cowan, A., Adkur, G., Killet, A., Brand, SL., Musa, MK., Meyer, J, Contextual factors influencing complex intervention research processes in care homes: a systematic review and framework synthesis. Age & Ageing, 2022. 51.

3. Staley, K., et al., What happens after James Lind Alliance Priority Setting Partnerships? A qualitative study of contexts, processes and impacts. Research involvement and engagement, 2020. 6(1): p. 41.

4. Deering, K., et al., Methodological procedures for priority setting mental health research: a systematic review summarising the methods, designs and frameworks involved with priority setting. Health Research Policy and Systems, 2023. 21(1): p. 64.

5. Davies, M., et al., Research collaboration with care home residents: a systematic review of public involvement approaches. Research Involvement and Engagement, 2025. 11(1): p. 49.

6. Prioritising Research through Engagement with older Adult Care Homes, P. Prioritising Research through Engagement with older Adult Care Homes (the PREACH project). 2025 [cited 2025 15 December]; Available from: https://www.carehomepriorities.com/.

7. James Lind Alliance, J. The James Lind Alliance Guidebook. 2021 March 2021 [cited 2025 26 November]; 10:[Available from: https://www.jla.nihr.ac.uk/jla-guidebook.

8. PREACH. Prioritising Research through Engagement with older Adult Care Homes (the PREACH project). 2024 [cited 2025 21 November]; Available from: https://www.carehomepriorities.com/.

9. Moore, D.C., et al., Factors associated with length of stay in care homes: a systematic review of international literature. Syst Rev, 2019. 8(1): p. 56.

10. Davies, M., Mathie, E., Irvine, L., Drury-Mulholland, M., Kelly, M., Killett, A., Care Home Activity Providers facilitating Public Involvement in research as meaningful activity for care home residents (CHAPPI) 2025.

11. Micklewright, K., et al., Activity provider-facilitated patient and public involvement with care home residents. Research Involvement and Engagement, 2024. 10(1): p. 7.

12. Killett, A., et al., Public involvement to enhance care home research; collaboration on a minimum data set for care homes. Health Expectations, 2025. 28(1): p. e70140.

13. Erlingsson, C. and P. Brysiewicz, A hands-on guide to doing content analysis. African journal of emergency medicine, 2017. 7(3): p. 93–99.

14. Graham, L., et al., What are the priorities for research of older people living in their own home, including those living with frailty? A systematic review and content analysis of studies reporting older people’s priorities and unmet needs. Age and Ageing, 2024. 53(1): p. afad232.

15. Irvine, L., et al., Data resource profile: the virtual international care homes trials archive (VICHTA). Int J Popul Data Sci, 2023. 8(6): p. 2161.

16. Irvine, L., et al., Protocol for the development of a repository of individual participant data from randomised controlled trials conducted in adult care homes (the Virtual International Care Homes Trials Archive (VICHTA)). Trials, 2021. 22(1): p. 157.

17. Tong, A., et al., Reporting guideline for priority setting of health research (REPRISE). BMC medical research methodology, 2019. 19(1): p. 243.

18. PREACH. Ideas for PREACH Sessions. 2025 [cited 2025 21 November]; Available from: https://padlet.com/lirvine11/ideas-for-preach-sessions-pn4kxp2n4hl51z9.

19. Cochrane, D.B., Research priority setting related to older adults: a scoping review to inform the Cochrane-Campbell Global Ageing Partnership work programme. 2022.

20. Devi, R., et al., The uncertainties and questions of care home residents, relatives and staff as a basis for evidence-based improvement and research. Age and Ageing, 2025. 54(5): p. afaf125.

21. Shepherd, V., F. Wood, and K. Hood, Establishing a set of research priorities in care homes for older people in the UK: a modified Delphi consensus study with care home staff. Age and Ageing, 2017. 46(2): p. 284–290.

22. Rayner, J.A., D. Fetherstonhaugh, and S. Cowen, Research priorities in residential aged care services: a statewide survey. Australasian journal on ageing, 2020. 39(1): p. 40–47.

23. Ali, M., et al., The virtual international stroke trials archive. Stroke, 2007. 38(6): p. 1905–1910.

24. Kelly, S., et al., Outcome measures from international older adult care home intervention research: a scoping review. Age and Ageing, 2023. 52(5): p. afad069.

25. Ruiz-Burga, E., et al., New horizons in improving research capacity in English care homes for older adults. Age and Ageing, 2025. 54(8): p. afaf211.

