## Supplementary material for "Prioritising Research through Engagement with older Adult Care Homes and potential for mapping to existing data sources: the PREACH study": Tables Figures & Appendices

PREACH Final Tables and Figures

### TABLES WITHIN MAIN PAPER

#### FIGURE 1: PREACH Framework

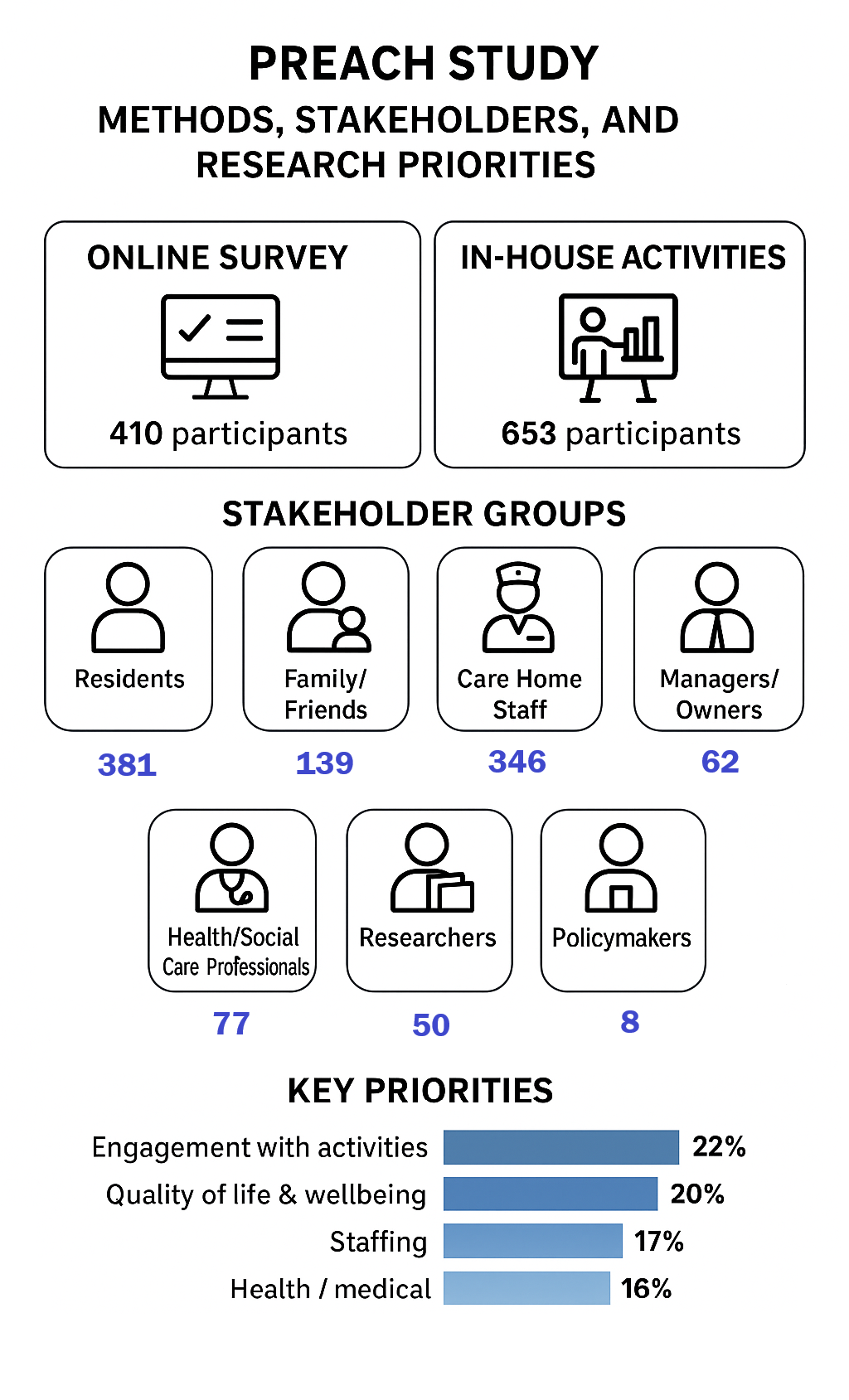

#### FIGURE 2: Frequency of idea (any mentions) by Stakeholder group

Frequency of ideas by Stakeholder group

|  | Care home resident | Care home staff member | Family or friend of care home resident | Care home owner or manager | Health care worker | Researcher | Social services worker | Policy & other |
| --- | --- | --- | --- | --- | --- | --- | --- | --- |
| Engagement with activities | 170 | 119 | 56 | 19 | 35 | 33 | 5 | 6 |
| Staffing | 80 | 131 | 52 | 43 | 51 | 36 | 3 | 6 |
| Quality of life and wellbeing | 112 | 161 | 61 | 22 | 26 | 19 | 1 | 2 |
| Health or medical | 92 | 94 | 42 | 38 | 63 | 30 | 5 | 6 |
| Quality of care & facilities | 74 | 89 | 49 | 18 | 35 | 15 | 8 | 4 |
| Exercise & mobility | 81 | 47 | 17 | 6 | 8 | 4 |  | 2 |
| Eating & drinking | 58 | 56 | 13 | 6 | 11 | 2 | 3 |  |
| Dementia | 15 | 45 | 25 | 14 | 15 | 14 |  | 2 |
| Funding & Finance | 32 | 35 | 26 | 14 | 5 | 8 |  | 4 |
| Choice & independence | 59 | 23 | 12 | 7 | 9 | 5 | 4 | 1 |
| Admissions/transitions | 34 | 19 | 18 | 10 | 14 | 7 | 4 | 1 |
| Medications | 23 | 37 | 12 | 14 | 12 | 4 | 2 | 2 |
| End of life care | 22 | 39 | 16 | 9 | 15 | 5 | 2 |  |
| Family involvement | 19 | 22 | 28 | 11 | 10 | 11 | 3 | 1 |
| Falls risk | 28 | 27 | 9 | 12 | 10 | 3 |  |  |
| Hospital visits | 31 | 15 | 9 | 10 | 18 | 1 | 2 | 2 |
| Technology | 12 | 20 | 7 | 7 | 5 | 9 |  | 3 |

#### TABLE 1: Examples of Activities mentioned

| **Types of activities mentioned** | **Examples** | **Number of mentions** |
| --- | --- | --- |
| **Exercise** | Strength building, embedding exercise into daily routine, “culture of movement”, social group exercise, swimming, dancing, chair yoga. | 105 |
| **Social connections** | Scheduled family nights, events in local community, baby yoga, contact with school children. | 98 |
| **Excursions / Days out** | Seaside trips, getting outside/into the garden more, access to gyms, going to cafes, organised day trips, walking groups. | 55 |
| **Cognitive / Learning** | Sign language class, dementia friends’ sessions, allowing chores, learning about medical conditions, emotional wellbeing sessions, grief counselling/support, lectures, reminiscence. | 33 |
| **Creative activities** | Decorating mobility aids, music therapy, gardening, games rooms, creating a craft shop, making art. | 29 |
| **Inclusive activities** | Activities for shy people, activities which accommodate for hearing/sight loss, specific activities for bed-ridden residents, sign-language/dementia choirs, cater for younger residents. | 24 |
| **Other** | Pets, importance of activity coordinators, religion-linked activities, importance of social mealtimes, beauty treatments, better use of technology including a “digital skills buddy scheme”. | 7 |

#### TABLE 2: Implications for policy, practice and research

**Resident-inclusive priority-setting is feasible at scale** and provides insights for researchers and funders

**Meaningful activity should be valued as core and resourced to enable this.** Residents consistently prioritised social connection, engagement and day-to-day autonomy over disease-focused concerns.

**Staff continuity and support are central to quality of life.** Workforce stability, protected time, and training directly affect residents’ ability to participate in meaningful activity.

Researchers planning care-home studies should **consider resident-identified priorities when selecting outcomes**, particularly measures of meaningful activity, social connection, autonomy and everyday wellbeing. Existing care-home datasets should be reused where appropriate, but data collection should also incorporate measures capable of capturing these domains.

The PREACH dataset provides an **openly available resource** that researchers can use to explore stakeholder-generated questions and apply alternative coding or analytical approaches.

#### FIGURE 3: Flowchart of stakeholder entry classification and viable research questions

*Processing 2013 stakeholder entries into 1528 original research ideas and 294 viable research questions, categorized by research design and potential for secondary dataset analysis*

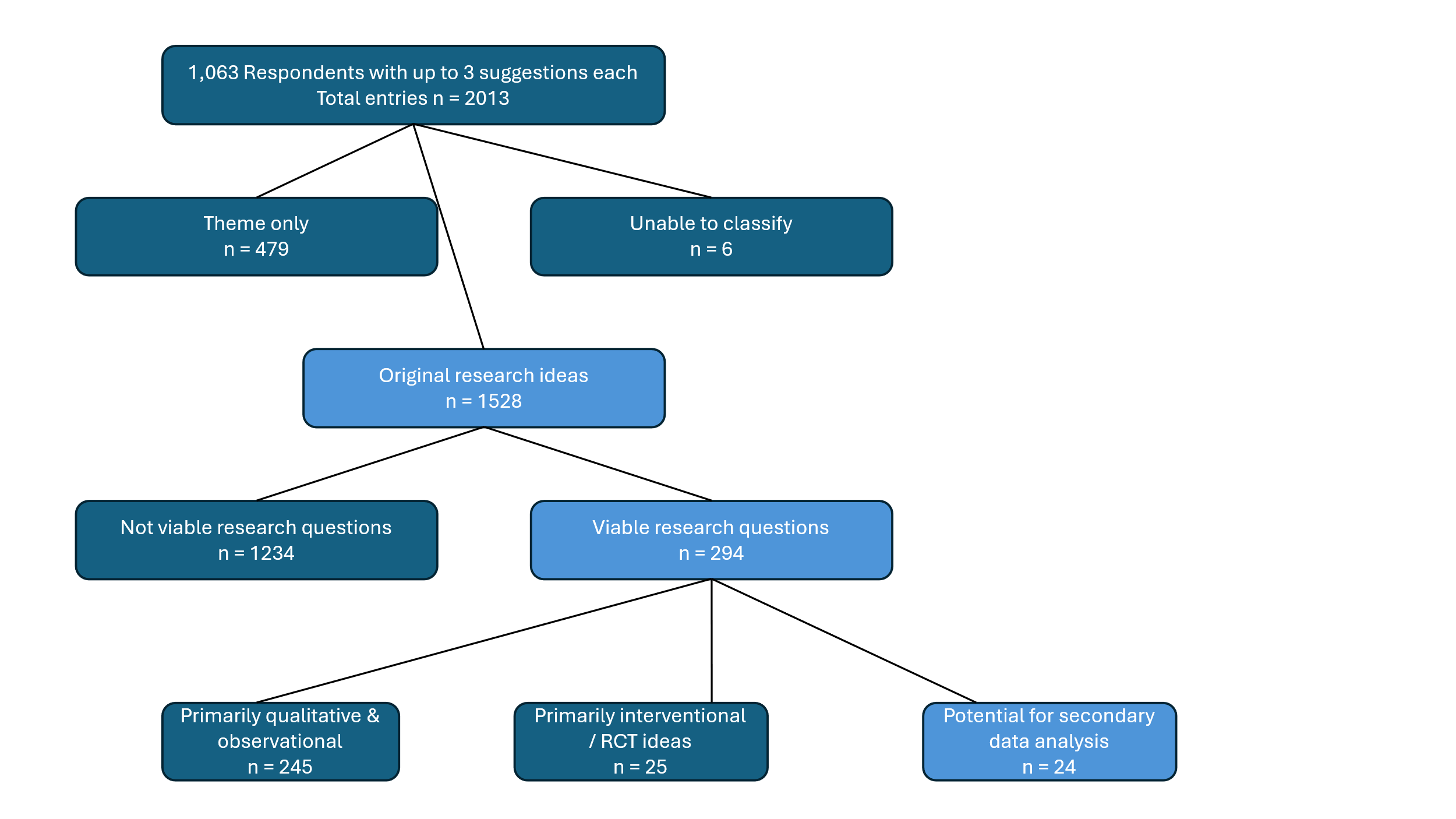

### Appendices

Appendices Table of Contents

##### A1 Online Survey

Participants were provided information, asked for informed consent, and then asked to indicate which stakeholder group they identified most closely with.

###### JISC ONLINE SURVEY SCREENSHOTS

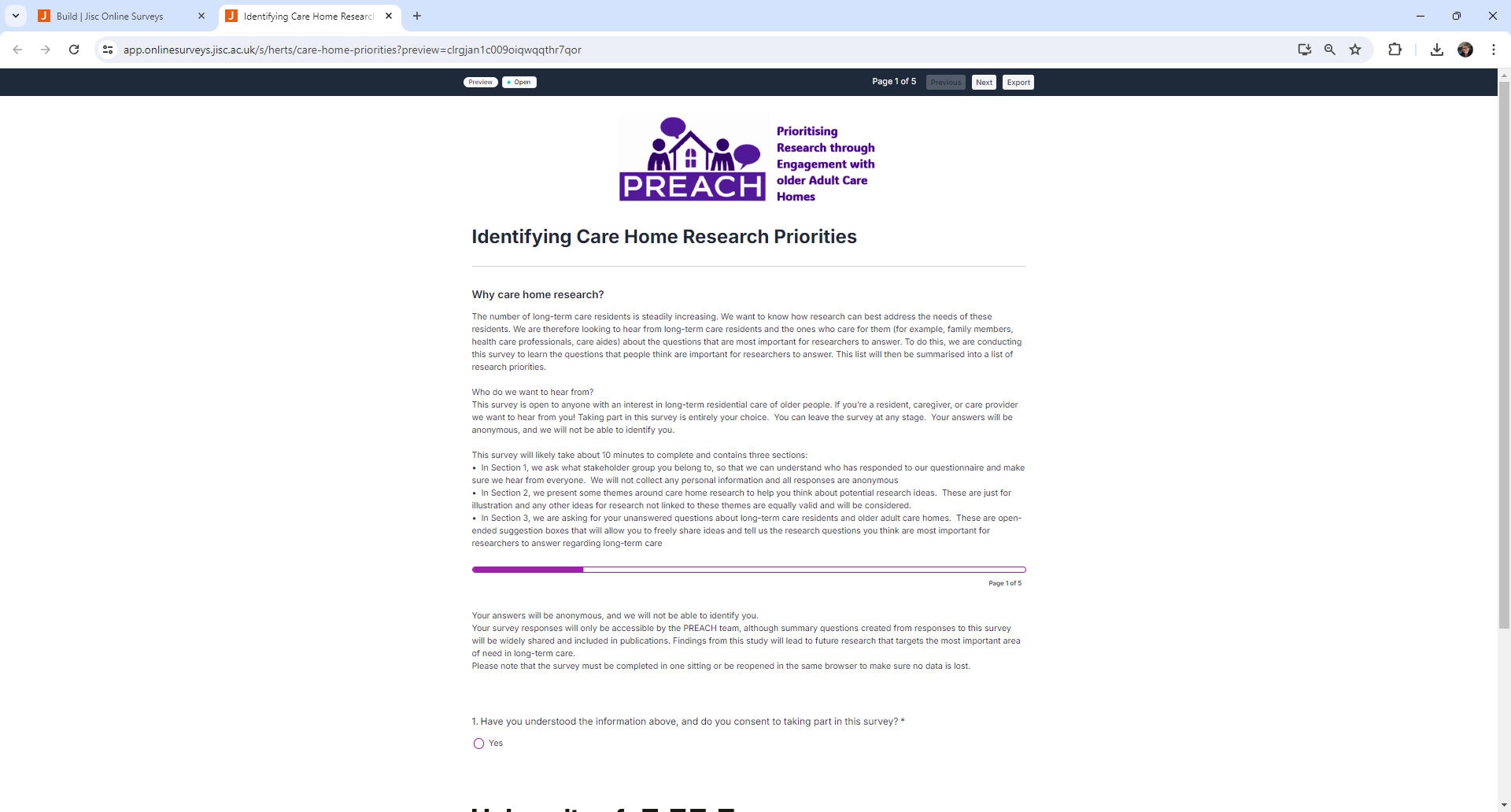

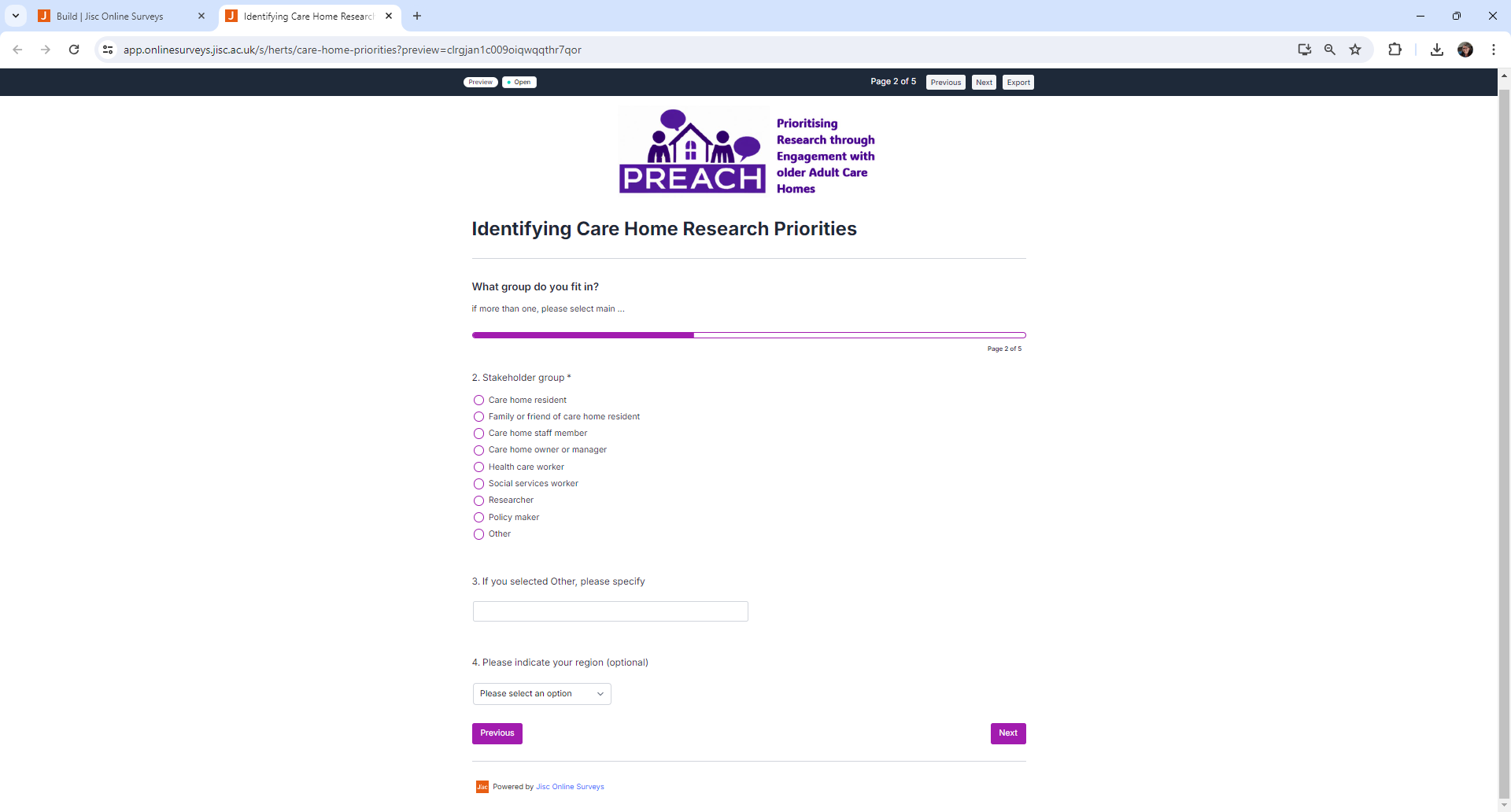

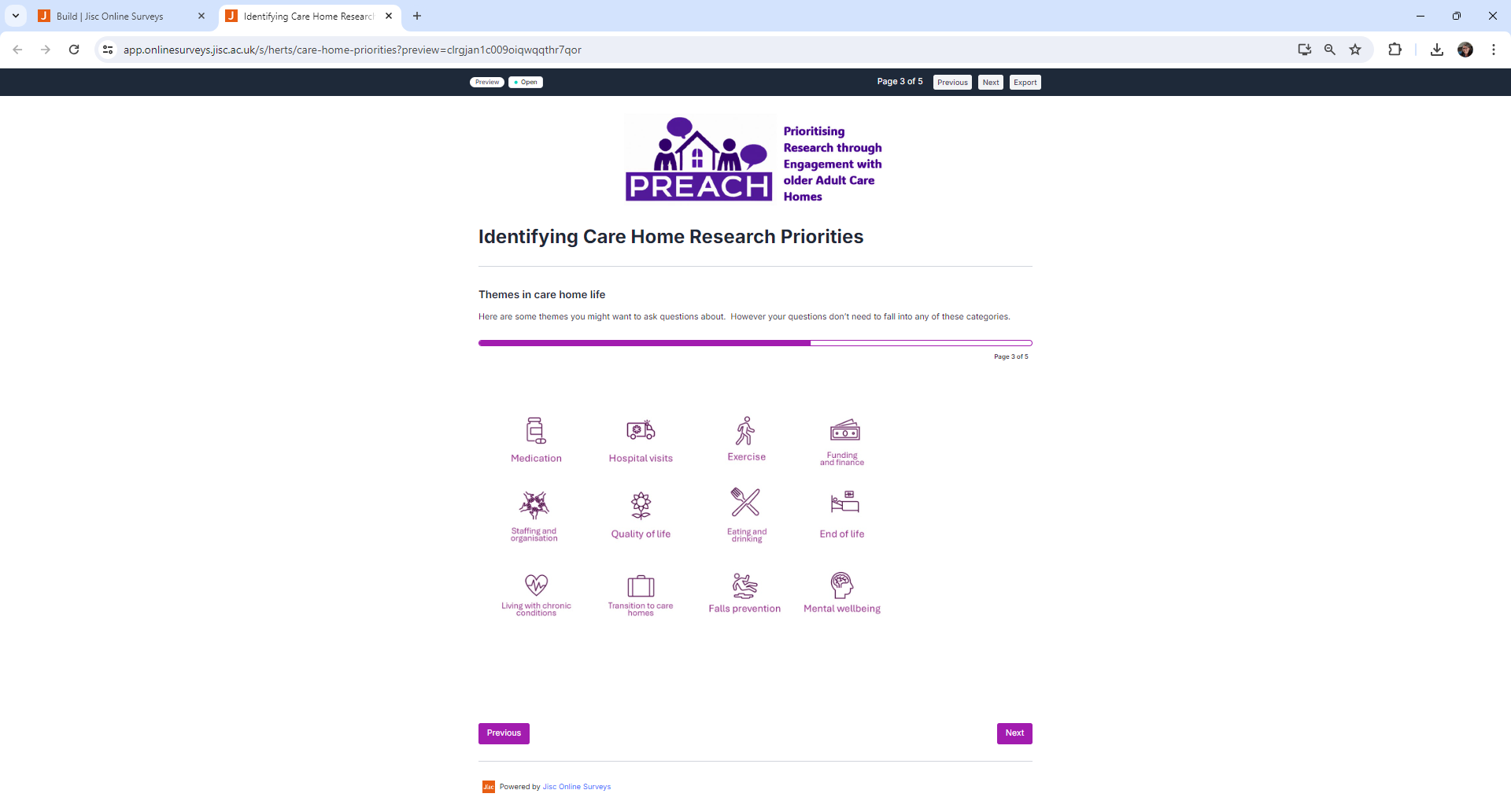

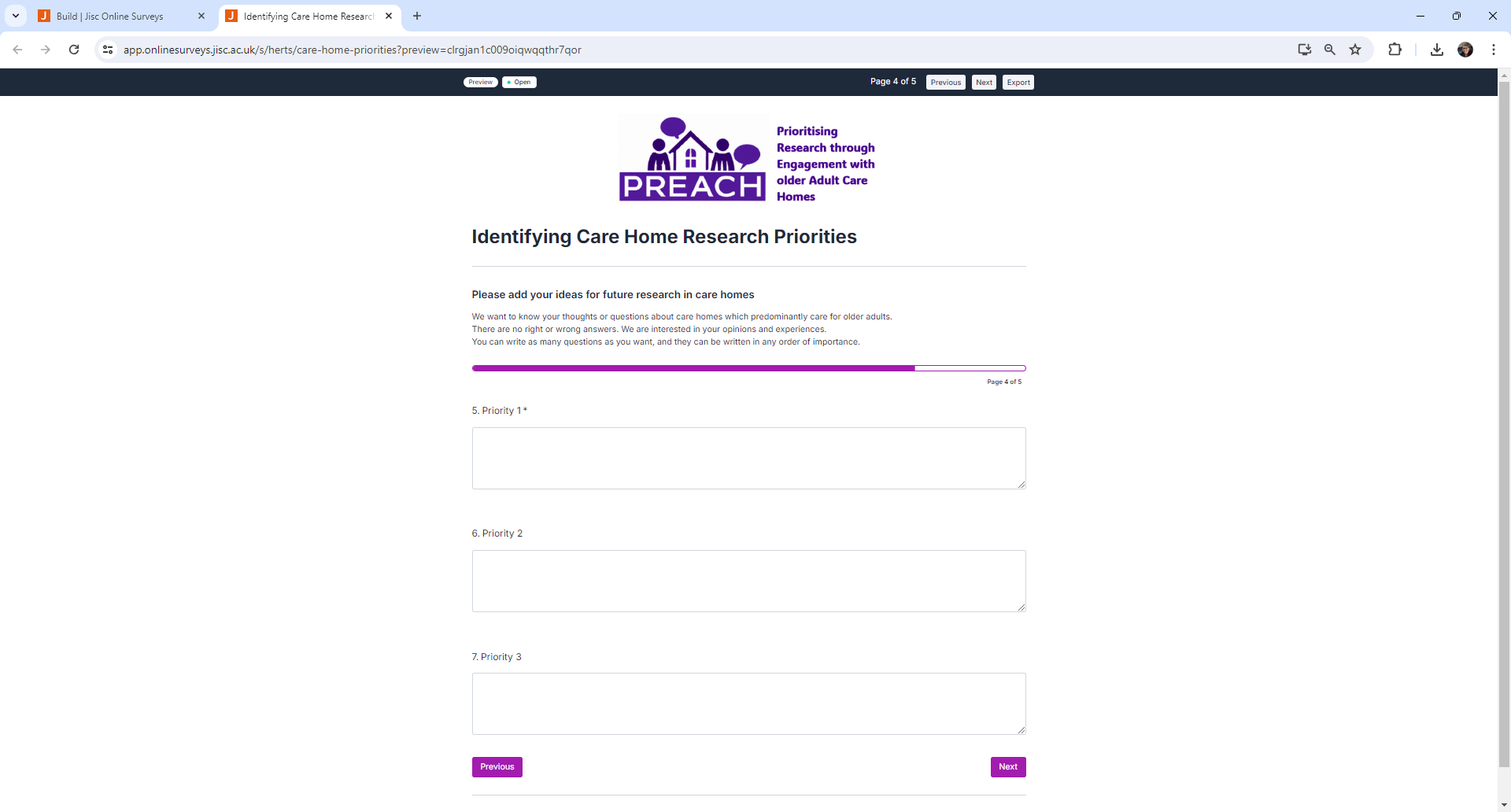

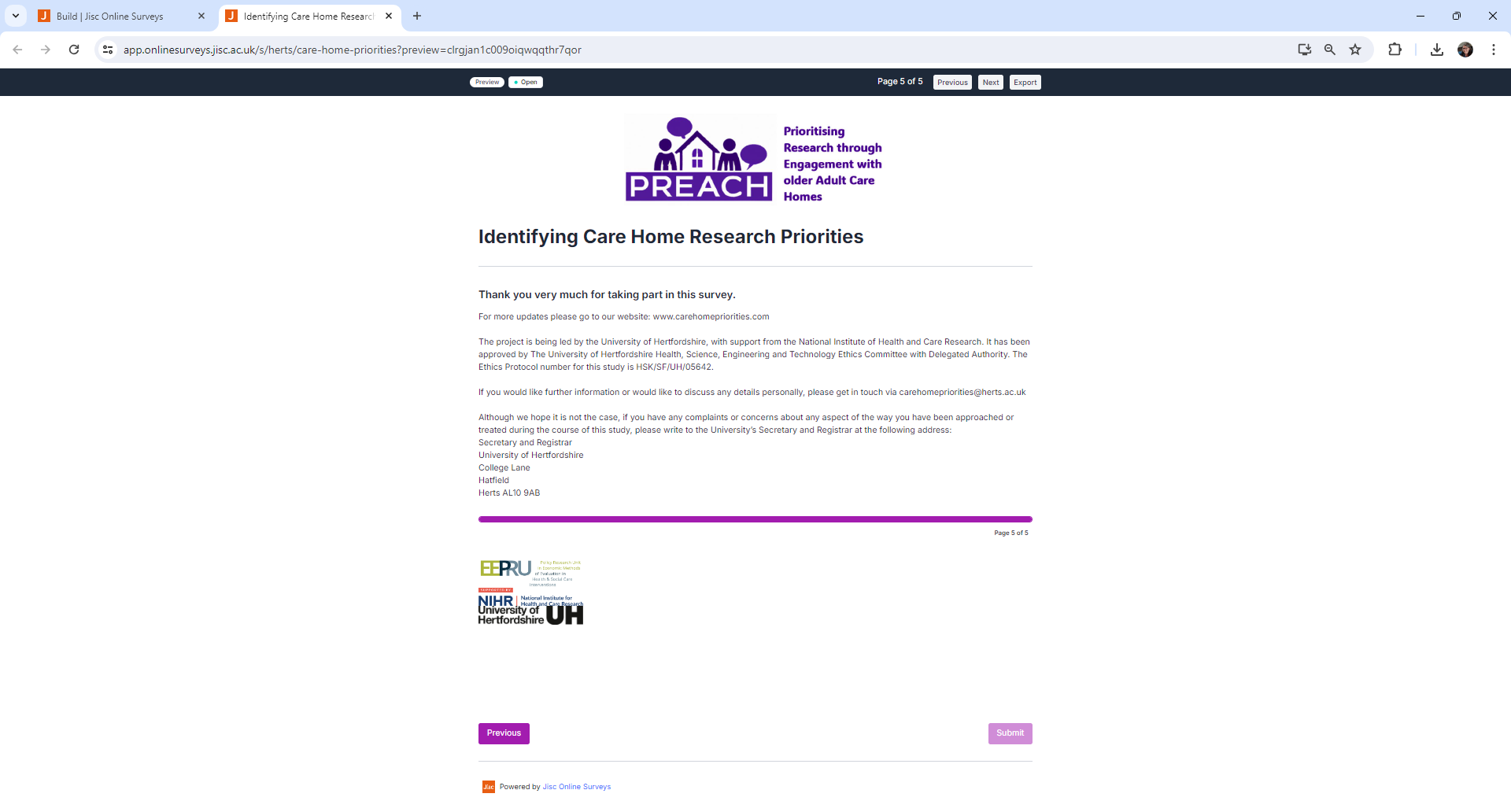

##### A2 Stakeholder groups and promotion activity

The online survey was promoted through: contacting care providers and advocacy organisations; creating blog posts on websites that stakeholders interact with; distributing information to research, profession-specific or special interest networks and forums (e.g., via newsletters); the study website; presentations and attendance to events aimed at stakeholder groups; and engagement with stakeholders via the National Institute for Health and Care Research (NIHR) Research Delivery Network (RDN), a government-funded organisation that supports the delivery of health and social care research

| Name of group/organization | Type of group/organization | Anticipated primary stakeholder type | Completed promotion activity |
| --- | --- | --- | --- |
| The Outstanding Society | Community Interest Company | Care home staff and managers | Distribution of flyer and information in member newsletter |
| British Geriatric Society (BGS) | Professional association | Healthcare professionals | Distribution of study information and link to survey via email |
| Royal College of Nursing (RCN) | Union and professional body | Healthcare professionals | Blog post (<https://www.rcn.org.uk/news-and-events/Blogs/why-nursing-voices-matter-in-shaping-care-home-research-230824>) |
| The Chartered Society of Physiotherapy – AGILE Professional Network | Union and professional body | Healthcare professionals | Distribution of study information and link to survey via email |
| Contact, Help, Advice and Information Network (CHAIN) | Online research support network | Researchers, healthcare professionals | Distribution of study information and link to survey via email |
| British Society of Gerontology (BSG) Special Interest Group | Professional organisation | Researchers, healthcare professionals, care home managers | Distribution of study information and survey link via newsletter |
| The National Institute for Health and Care Research (NIHR) Economic Methods of Evaluation of Health and Care Interventions Policy Research Unit | Research collaboration | Researchers, policy makers | Social media posts |

##### A3 Promotional materials

###### A3a Poster

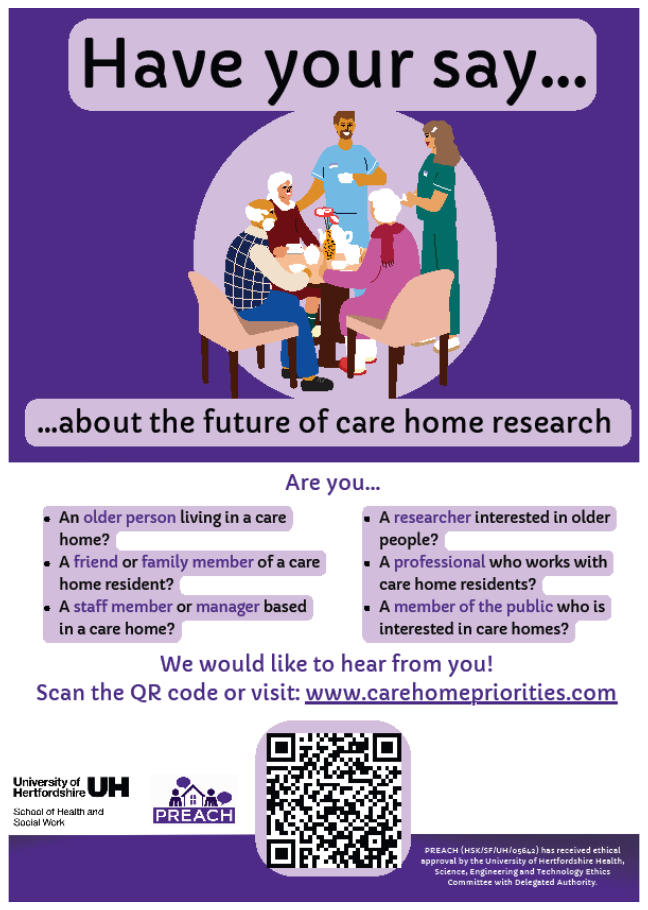

###### A3b Activity packs

Available to download from study website: <https://www.carehomepriorities.com/activity-packs-to-download>

These were activities and suggestions about how to explore residents’ views on priorities for care home research. We did not need a set number of people to take part. We were keen to hear from anyone who is interested and comfortable with sharing their opinions. Homes are not expected to do every activity - we appreciate any feedback you can give. If residents’ family or friends were visiting they were welcome to take part too. We were keen to hear from staff members, volunteers and anyone else who supported residents.

Idea 1: Icebreaker (Quiz) – a true or false quiz about research breakthroughs and care home facts. It is nine questions long and has printable A4 “Yes/No/Not sure” prompt cards included.

Idea 2: Icebreaker (Tile matching game) - a simple game involving making pairs of matching images.

Idea 3: Answer cards – printable cards that can be cut up and given to participants so that they can write down their ideas about research. These could be paired with a suggestions box or used in activity sessions.

Idea 4: Discussion prompts – ideas to help facilitate a discussion around care home research in the news.

Idea 5: Priorities poster – a printable poster to allow participants to “vote” for research topics.

Idea 6: Prompt cards – A4 prompt cards with pictures and large text to help facilitate activities and discussions.

###### A3c Suggestion cards

Delivered to each bedroom & used in group discussions with residents

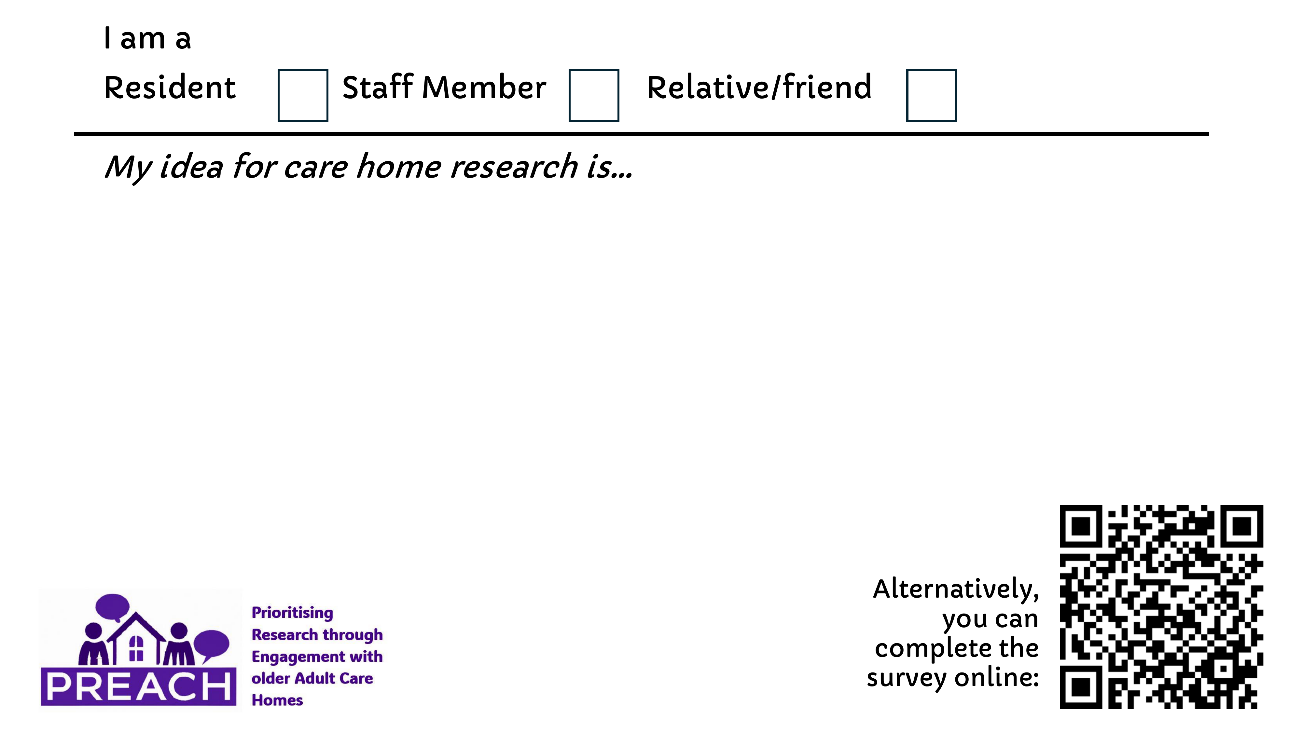

###### A3d Bifold (resident) Flyer &

Delivered to each bedroom & used in group discussions with residents

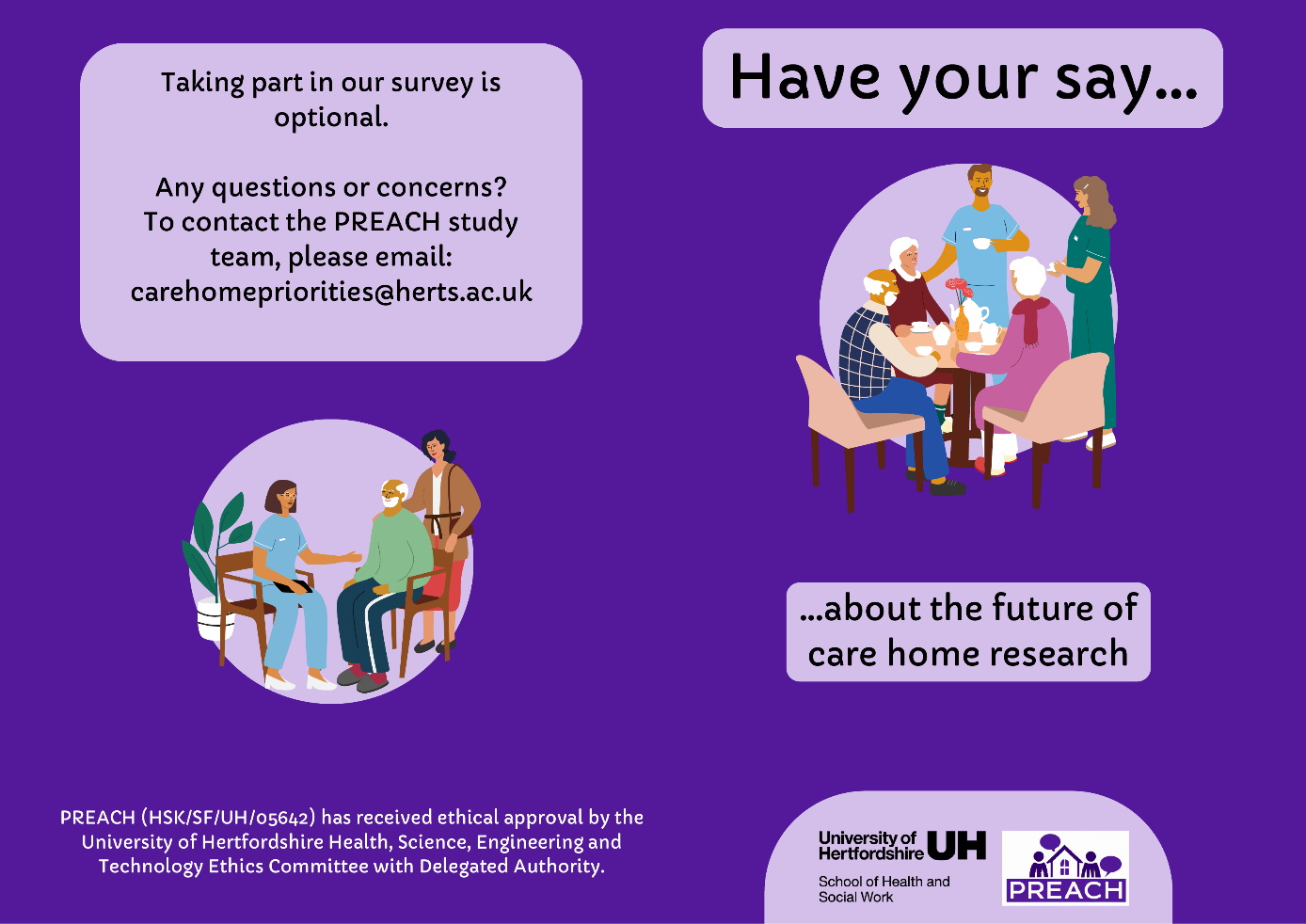

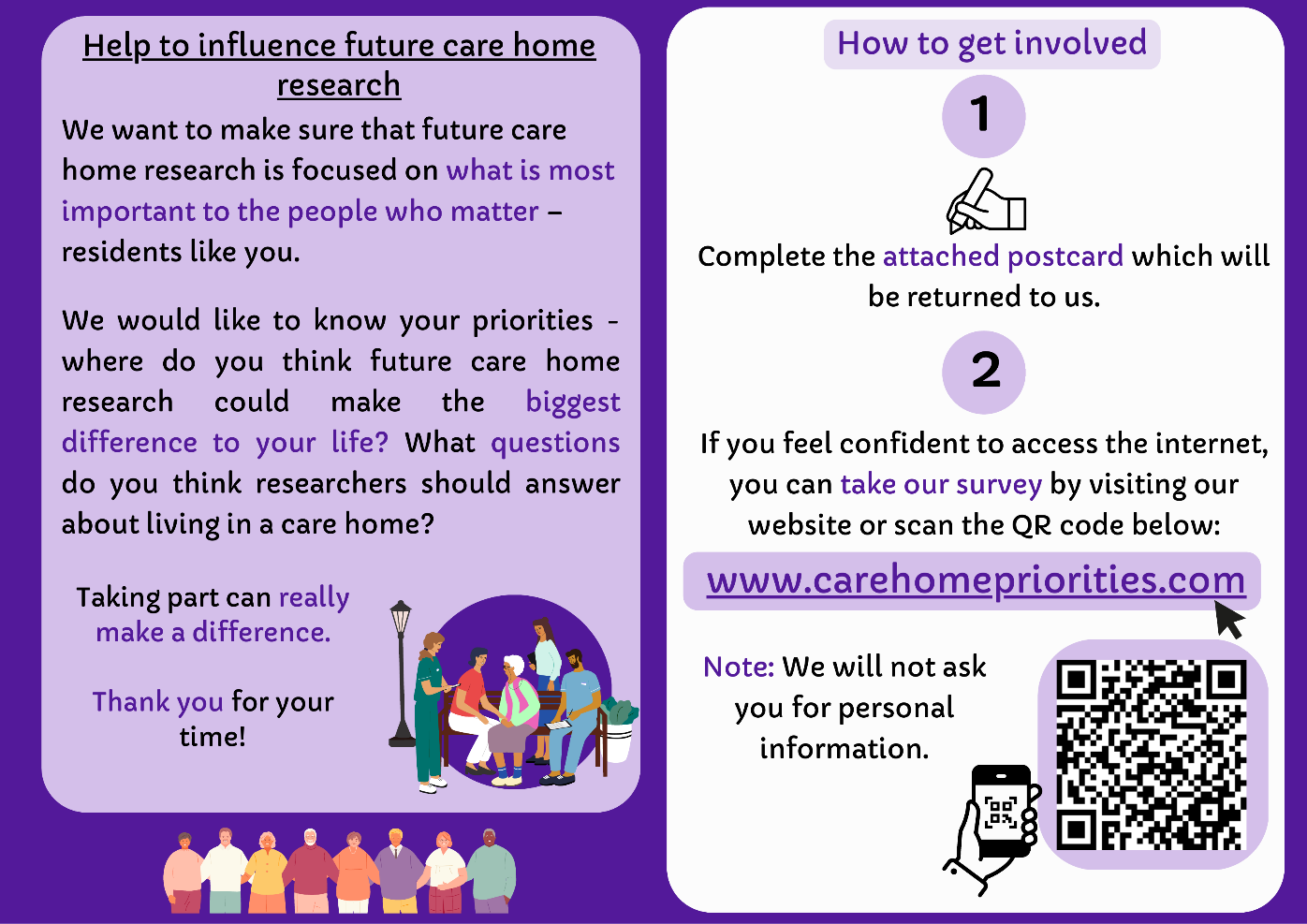

###### A3e Trifold flyer – staff and visitors

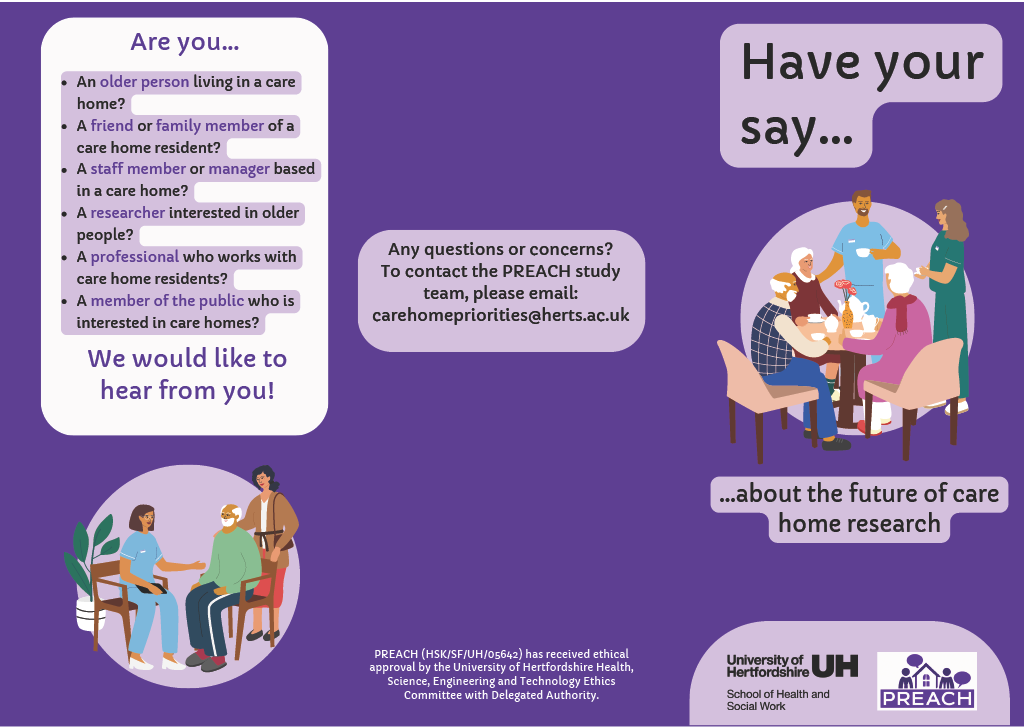

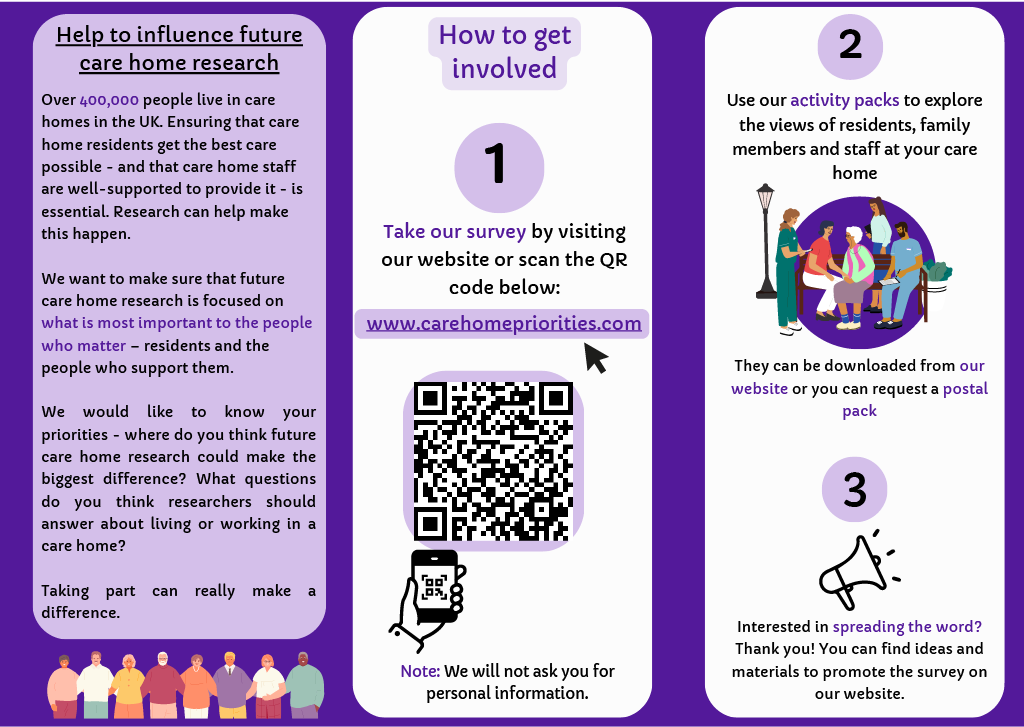

##### A4 Qualitative Content Analysis Methodology

Survey and postal responses were combined and analysed in Excel. Qualitative content analysis was used to systematically code individual contributions and generate themes [16]. Themes from a recent review and content analysis of priority setting work with older people [17] were used for the initial coding framework. Where responses contained multiple distinct ideas, each idea was coded separately, allowing individual submissions to contribute to more than one code and theme. Inductive analysis was conducted and new codes were added when needed by KM, Emerging codes and themes were discussed regularly by KM and LI., and any uncertainties regarding code allocation or theme definitions were resolved iteratively through discussion and consensus. Coding continued until no substantively new priority areas emerged and the coding framework remained stable across additional responses. An analysis log was maintained throughout coding to track uncertainties, any changes to coding and decisions made. Duplicate ideas were not removed, as repeated mentions were considered indicative of shared importance. Instead, prioritisation was based on the frequency with which each idea was raised, with those mentioned most often given higher priority. After completion of coding, frequencies were calculated by summing the number of responses assigned to each final theme. Where responses contributed to multiple themes, each coded theme contributed independently to the overall frequency counts. The distribution of themes across stakeholder groups was also analysed. PREACH sought to capture stakeholder-generated ideas without constraining responses. Frequency should therefore be interpreted as indicating how commonly topics were raised rather than their relative importance. Due to our metrics-based design (rather than consensus-based), a second-stage prioritisation exercise was not conducted. Coding was undertaken using Microsoft Excel, which provided a transparent audit trail and facilitated management of the large volume of short-text responses. Given the brevity of submissions and emphasis on descriptive content analysis, specialist qualitative software was not considered necessary.

Qualitative content analysis was selected because the study sought to systematically describe the frequency and content of stakeholder-generated research priorities across a large dataset of predominantly brief textual responses. Unlike reflexive thematic analysis, which aims to generate rich interpretive understanding, content analysis is well suited to organising large numbers of concise responses into descriptive categories while allowing quantitative summary of theme frequencies.

PREACH deliberately focused on generating priorities rather than producing a ranked consensus list. As all submissions were anonymous, this prevented follow-up or clarification of ambiguous suggestions. In line with data minimisation principles, we did not collect demographic data which limits our ability to assess equity and understand whose voices were most/least represented.

##### A5 Mapping to Existing data Methodology

**1. Question Viability & Operationalisation:**

Submissions were first filtered to identify **"viable research questions"**. We defined a viable question as an idea articulating a testable relationship between an exposure (e.g., intervention, organisational attribute) and an outcome (e.g., quality of life, fall rates), or a hypothesis amenable to structured empirical inquiry. General descriptive statements, isolated complaints, or broad expressions of preference (e.g., *"the food should be better"*) were classified as non-viable research questions for secondary quantitative analysis, though preserved within qualitative thematic findings.

**2. Mapping Classification Criteria:**

To assess whether stakeholder priorities could be investigated using existing data, each viable research question was compared against the variables available within VICHTA. Questions were classified into one of three categories:

- *Addressable* – the required exposure(s), outcome(s) and study population were available within VICHTA, such that the question could reasonably be examined using secondary analysis.
- *Partially addressable* – some, but not all, required variables were available, meaning that only part of the research question could be explored, or a proxy measure would be required.
- *Not addressable* – one or more essential concepts were absent from VICHTA, preventing meaningful investigation using the available data.

Mapping inevitably involved a degree of judgement because stakeholder-generated priorities were often broad or expressed in everyday language rather than as fully specified research questions. To minimise subjectivity, classification criteria were agreed before mapping commenced.

Initial mapping was undertaken by XX and independently reviewed by XX. Any uncertainties or disagreements were resolved through discussion with the wider research team until consensus was reached. While minor differences in classification might reasonably occur between reviewers, we anticipate that the overall conclusions regarding the limited alignment between stakeholder priorities and existing datasets would remain unchanged.

**3. Results**

Of the 2,013 total stakeholder entries, 1,528 represented original research ideas. Content and feasibility analysis identified 294 viable research questions (19.2% of original ideas). These were categorised by suitable research design: qualitative designs (n=245), primary randomised trial designs (n=25, Appendix A13), and potential secondary dataset analysis (n=24, Appendix A14).

When mapped against the VICHTA dataset:

- *Fully Addressable*: No stakeholder-generated question was completely addressable using existing VICHTA variables alone.
- *Partially Addressable* (n=24, 8.2% of viable questions): Analysis was limited to clinical and health-service constructs where proxy measures existed. These clustered in four clinical domains:
  1. Falls and physical function (e.g., falls incidence vs. exercise availability).
  2. Medication management (e.g., polypharmacy metrics and proxy QoL measures).
  3. Hospital admissions & acute care reliance.
  4. Cognitive status / dementia progression.
- *Not Addressable* (n=270; 91.8% of viable questions): The vast majority of viable questions could not be addressed. Infeasibility was primarily driven by missing exposure variables (e.g., staff turnover rates, activity participation logs, relational care practices) or missing experiential outcome measures (e.g., sense of autonomy, loneliness, mealtime satisfaction).

| **Stakeholder priority** | **Research question** | **VICHTA variables** | **Classification** | **Reason** |
| --- | --- | --- | --- | --- |
| More meaningful activities | Does participation in organised activities improve quality of life? | Activity participation ❌; DEMQOL ✓ | Partially addressable | Outcome available, routine activity dose/type exposure absent |
| Staff continuity | Does lower staff turnover improve resident QoL? | Staff turnover ❌ | Not addressable | Exposure unavailable |
| Falls prevention | Does exercise reduce falls? | Falls ✓; intervention ✓ | Partially addressable | Clinical trial variables present, but long-term maintenance data lacking |
| Mealtime experience | Does social/communal dining improve hydration and QoL? | Dining environment  Weight / QoL | Not addressable | Environmental and relational care process variables absent |

**4. Discussion**

Inability to map not a negative result or failure, but a major structural finding. Trial archives like VICHTA were historically built around disease-oriented RCTs (measuring falls, cognition, medication, hospitalisations). The lack of alignment reflects a systemic bias in trial design. Researchers routinely collect data on clinical endpoints, whereas residents prioritize relational care, staff stability, daily autonomy, and meaningful activity. Argue for standardised collection of Resident-Reported Outcome Measures (RROMs) and routine recording of organizational variables (such as staffing ratios and activity provision) in future primary trial datasets so secondary archives can eventually answer these vital questions.

##### A6 REPRISE checklist

| **No** | **Item** | **PREACH** |
| --- | --- | --- |
| **A** | **Context and scope** |  |
| 1 | Define geographical scope | UK  (can accept global responses but all VICHTA data is from UK so far) |
| 2 | Define health area, field, focus | Older adults living in residential or nursing homes |
| 3 | Define the intended beneficiaries | Care home residents  Care home staff and managers  Family carers  Local authorities/commissioners  Health and social care professionals  Regulators  Researchers |
| 4 | Define the target audience of the priorities | Priorities are intended to inform researchers, funders, policymakers and care-home organisations |
| 5 | Identify the research area | Care home research/long term care research |
| 6 | Identify the type of research questions | Priorities for care home research; may relate to health and wellbeing (physical and/or psychological), care/service provision, funding, or other related topics. |
| 7 | Define the time frame | No plans to revise or update following project’s conclusion |
| **B** | **Governance and team** |  |
| 8 | Describe the selection and structure of the leadership and management team | Small team of two researchers led the project; both have previous experience of research relating to care homes/older people. The study team reported to funders, XXX |
| 9 | Describe the characteristics of the team | Both researchers were based in the UK (University of XXX). The lead researcher is a health economist by background, while the other researcher is an allied health professional who works with older people and has experience of Patient and Public Involvement (PPI) in research. Both researchers have previously been involved in care home research. |
| 10 | Describe any training or experience relevant to conducting priority setting | XX has expertise in qualitative methods including thematic analysis. XX has experience in survey design, nominal group analysis, ethnography and thematic analysis using NVivo software |
| **C** | **Framework for priority setting** | |
| 11 | State the framework used (if any) | Responses initially coded to domains based on Graham et al.’s priority setting work with older people living in their own homes (doi: 10.1093/ageing/afad232), though the team anticipate responses that will fall outside of these (for example, responses specific to care provision within residential or nursing homes). As such, it is likely additional domains will be added as analysis continues. |
| **D** | **Stakeholders or participants** | |
| 12 | Define the inclusion criteria for stakeholders involved in priority-setting | Care home residents  Care home staff and managers/care providers  Care provider advocacy organisations  Family carers  Local authorities/commissioners  Health and social care professionals  Regulators  Researchers  Demographic characteristics were not collected |
| 13 | State the strategy or method for identifying and engaging stakeholders | Identifying stakeholders: utilising knowledge from previous research; searching online; word of mouth; advice from stakeholder representatives (PPI) and the Advisory Group.  Engaging stakeholders: creation of activity packs to be used with care home residents; contacting care providers and advocacy organisations; creating blog posts on websites that stakeholders interact with; distributing information to research or special interest networks and forums; invitations to participate distributed via newsletters of stakeholder groups/organisations; study website (with additional resources inc. promotional guide and materials available for visitors); presentations and attendance to events aimed at stakeholder groups; engagement with stakeholders via Clinical Research Network support. |
| 14 | Indicate the number of participants and/or organizations involved | 1,063 stakeholders contributed to the study, as detailed below:   - Residents – 381 - Care home staff – 346 - Family & friends – 139 - Health/social care professionals – 77 - Care home manager/owners – 62 - Researchers – 50 - Policymakers/others – 8 |
| 15 | Describe the characteristics of stakeholders | Stakeholders should have personal or professional experience of, or interest in, care homes. |
| 16 | State if reimbursement for participation was provided | N/A per survey participant  An incentive structure was designed to recognise institutional effort without introducing individual bias. Participating homes were entered into a prize draw for £50 Love2Shop vouchers, while RDN resources were utilised to offset the delivery costs for frontline sites. |
| **E** | **Identification and collection of research priorities** | |
| 17 | Describe methods for collecting initial priorities | Online survey via JISC platform and response cards returned from in-person resident engagement. Priorities from previous relevant priority setting work/research also included. |
| 18 | Describe methods for collating and categorizing priorities | Data was managed via NVivo 14 and analysed in Microsoft Excel. Duplicate retention strategy was employed, as frequency serves as a proxy for community urgency, whereas deduplication may flatten the perceived significance of widespread concerns |
| 19 | Describe methods and reasons for modifying (removing, adding, reframing) priorities | Intentionally broad phrasing:  Online survey “Please add your ideas for care home research” – followed by three text boxes. Each text box coded separately.    Suggestion card “My idea for care home research is…” |
| 20 | Describe methods for refining or translating priorities into research topics or questions | Reviewed by analysis team (XX) |
| 21 | Describe methods for checking whether research questions or topics have been answered | Priorities were not assessed against existing literature/evidence, because the purpose was to assess their alignment with existing datasets  Mapping undertaken by XX with support from XX |
| 22 | Describe number of research questions or topics | The team translated 2,013 raw ideas into 294 viable research questions. Only 2% of stakeholder-prioritised topics were directly addressable using existing secondary datasets within VICHTA |
| **F** | **Prioritization of research topics/questions** | |
| 23 | Describe methods and criteria for prioritizing research topics or questions | Ranking was determined by the frequency of mentions across all stakeholder groups. This model prioritised shared importance, allowing the concerns of residents and staff (who formed the bulk of the respondents) to naturally lead the agenda.  One-off suggestion card/survey entry – no facility to come back to participants for consensus or ranking |
| 24 | State the method or threshold for excluding research topics/questions | To maximise the utility of the findings, no topics were excluded. The study preserved the full 51-theme dataset as a public asset, ensuring that even niche research uncertainties remain accessible for specialised secondary analysis |
| **G** | **Output** |  |
| 25 | State the approach to formulating the research priorities | The findings revealed a clear hierarchy of stakeholder concerns:   1. **Activities (22%):** Meaningful engagement, social connection, and continued learning. 2. **Quality of Life and Wellbeing (20%):** Autonomy, resident choice, and independence. 3. **Staffing (17%):** Retention, training, and safe staffing levels. 4. **Health and Medical (16%):** Access to GPs and prevention of unnecessary hospitalisations. |
| **H** | **Evaluation and feedback** | |
| 26 | Describe how the process of prioritization was evaluated | The team utilised facilitator feedback forms and an anonymous study [Padlet](https://padlet.com/lirvine11/ideas-for-preach-sessions-pn4kxp2n4hl51z9) to evaluate the engagement model. No formal process evaluation was performed. |
| 27 | Describe how priorities were fed back to stakeholders and/or to the public; and how feedback (if received) was addressed and integrated | Dissemination included a "Headline Results" [newsletter](https://www.carehomepriorities.com/_files/ugd/3ce93d_cb114241e30d460495f0beb1150ec760.pdf), the public website (www.carehomepriorities.com), and XXX webinars. |
| **I** | **Implementation** |  |
| 28 | Outline the strategy or action plans for implementing priorities | The deidentified dataset is available in Excel format for public re-analysis. The priorities have been formally presented to NIHR commissioners to influence future funding calls, particularly for the 245 questions requiring primary data collection |
| 29 | Describe plans, strategies, or suggestions to evaluate impact | Impact will be tracked through citations and downloads of the PREACH database, and the development of secondary data projects addressing the questions mapped to the VICHTA trials archive. Further evaluation impact is outside the scope of the study. |
| **J** | **Funding and conflict of interest** | |
| 30 | State sources of funding | This study was funded by XXXX |
| 31 | Declare any conflicts or competing interests | No conflicts or competing interests were declared by the research team |

Research protocol available on project website: <https://www.carehomepriorities.com/about-preach>

##### A7 Number of respondents per data collection method

| **Stakeholder group** | **In-person** | **Online** | **Grand Total** |
| --- | --- | --- | --- |
| Care home staff member | 223 | 123 | 346 |
| Health care worker |  | 70 | 70 |
| Care home owner or manager |  | 62 | 62 |
| Family or friend of care home resident | 77 | 62 | 139 |
| Researcher |  | 50 | 50 |
| Care home resident | 353 | 28 | 381 |
| Policy & other |  | 8 | 8 |
| Social services worker |  | 7 | 7 |
| **Grand Total** | **653** | **410** | **1063** |

##### A8a Care home characteristics

| **Care home characteristic** | | **N PREACH** | **% PREACH sample** | **% National** |
| --- | --- | --- | --- | --- |
| Type of provider | Privately owned | 39 | 78.0 | 84.3 |
|  | Voluntary / not for profit | 8 | 16.0 | 13.3 |
|  | Local authority | 3 | 6.0 | 2.4 |
| Size of organisation | Large multi-chain (>51) | 3 | 6.0 | 19.3 |
|  | Medium-sized (2-50) | 24 | 48.0 | 19.3 |
|  | Single owner | 20 | 40.0 | 61.4 |
|  | Local Authority | 3 | 6.0 | - |
| Geographical region | Urban | 32 | 64.0 | unknown |
|  | Rural | 18 | 36.0 | unknown |
| Type of Service | Nursing | 20 | 40.0 | 28.4 |
|  | Residential only | 30 | 60.0 | 49.6 |
| No. beds | <30 beds | 12 | 24.0 | 59.2 |
|  | 31-60 beds | 27 | 54.0 | 29.1 |
|  | 61+ beds | 11 | 22.0 | 11.7 |
| CQC Rating | Outstanding | 6 | 12.5 | 4.4 |
|  | Good | 37 | 77.1 | 78 |
|  | Requires Improvement | 5 | 10.4 | 16.1 |
|  | N/A (newly opened home) | 2 | 4.2 | 1.4 |
| Range of fees (Self-fund residential care per week) | Range: £850 - £2411 | 33 | 66.0 | unknown |
|  | Not publicly available | 14 | 28.0 | unknown |
|  | Local Authority | 3 | 6.0 | unknown |

*Publicly available on carehome.co.uk, accessed June 2025

*Statutory data published by CQC, as reported in Hanratty 2023

*Hanratty B, Wolters AT, Towers AM, Spilsbury K, Meyer J, Killett A, Jones L, Gordon A, Burton JK, Akdur G, Irvine L. Data Collection in Care Homes for Older Adults: A National Survey in England. Journal of Long-Term Care. 2023 Jan 1.*

##### A8b Map of included care homes

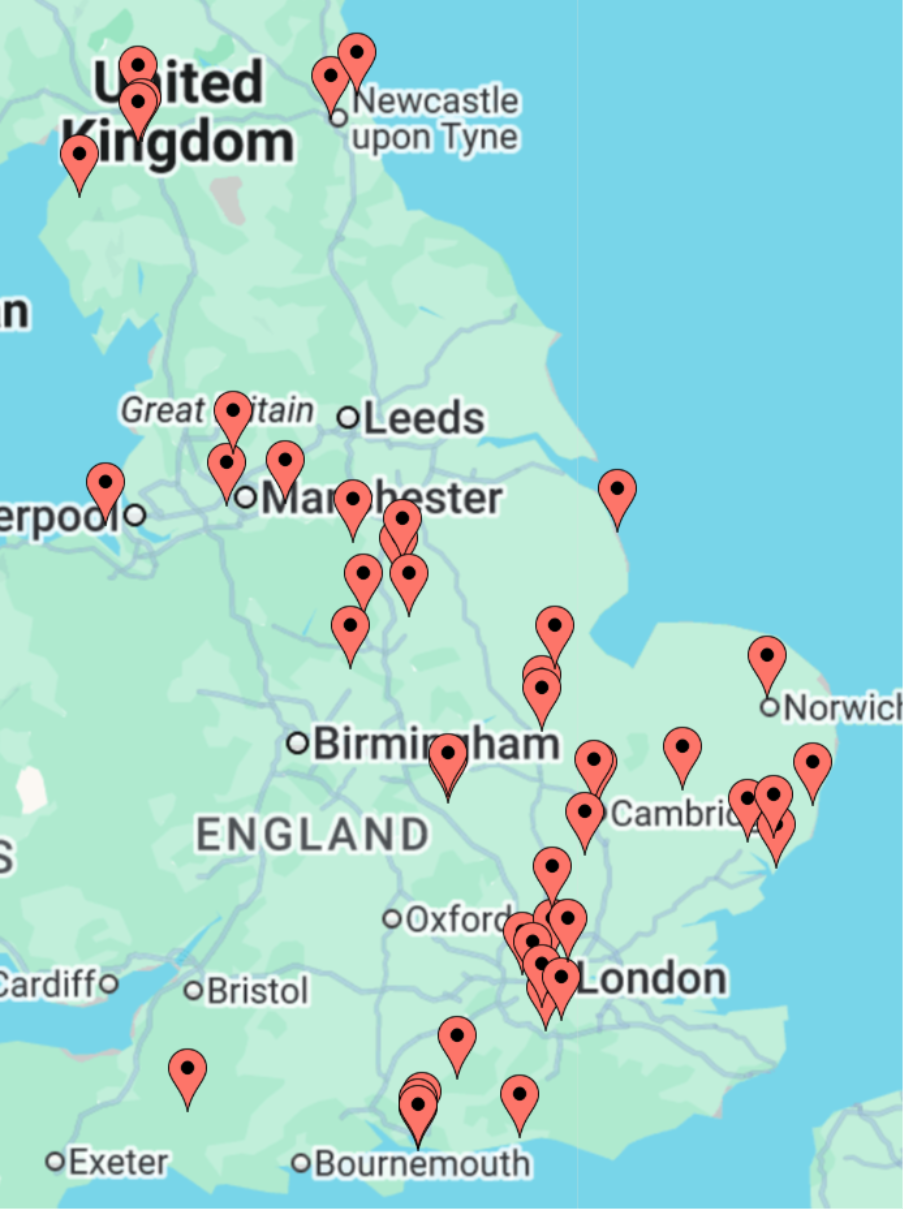

##### A9 “Headline results” Newsletter to Care homes

Downloadable on study website: <https://www.carehomepriorities.com/headline-results>

##### A10 Full list of research themes and mentions per stakeholder group

Readers are encouraged to use the raw data in line with their interests. The dataset is provided as an Excel file, allowing users to filter responses identified by the study team or to apply new coding and reclassifications to suit different analytical frameworks.

|  | **Stakeholder group** | | | | | | | |  |
| --- | --- | --- | --- | --- | --- | --- | --- | --- | --- |
| **Theme** | **Care home owner or manager** | **Care home resident** | **Care home staff member** | **Family or friend of resident** | **Health care worker** | **Policy & other** | **Researcher** | **Social services worker** | **Total mentions** |
| ID | 149 | 574 | 675 | 263 | 188 | 23 | 120 | 21 | 2013 |
| **Engagement with activities** | 19 | 175 | 120 | 56 | 35 | 6 | 33 | 5 | 443 |
| Exercise & mobility | 6 | 81 | 47 | 17 | 8 | 2 | 4 |  | 165 |
| Intergenerational links | 1 | 9 | 4 | 2 |  |  | 4 | 1 | 21 |
| Music / Sound / Noise | 1 | 16 | 10 | 2 | 2 | 3 |  |  | 34 |
| Getting outside & community links | 2 | 47 | 19 | 9 | 8 | 1 | 8 | 2 | 96 |
| **Quality of life and wellbeing** | 22 | 114 | 161 | 61 | 26 | 2 | 19 | 1 | 406 |
| Resident choice, autonomy and independence | 7 | 59 | 23 | 12 | 9 | 1 | 5 | 4 | 120 |
| Social connection & Loneliness | 3 | 34 | 19 | 16 | 6 |  | 5 | 2 | 85 |
| Managing distress | 1 | 1 | 4 | 3 |  |  | 1 |  | 10 |
| **Staffing** | 43 | 80 | 131 | 52 | 51 | 6 | 36 | 3 | 402 |
| Staff training | 12 | 10 | 29 | 6 | 17 | 3 | 5 | 1 | 83 |
| Staff wellbeing, pay, retention | 10 | 12 | 30 | 7 | 7 | 1 | 6 |  | 73 |
| Staffing levels | 4 | 8 | 25 | 7 | 8 |  | 8 |  | 60 |
| Staff recruitment | 3 | 1 | 5 | 1 | 4 |  | 10 |  | 24 |
| **Health or medical** | 40 | 95 | 96 | 42 | 64 | 6 | 30 | 5 | 378 |
| Dementia | 14 | 15 | 45 | 25 | 15 | 2 | 14 |  | 130 |
| Medications | 14 | 23 | 37 | 14 | 12 | 2 | 4 | 2 | 108 |
| End of life care | 9 | 22 | 39 | 16 | 15 |  | 5 | 2 | 108 |
| Falls risk | 12 | 28 | 27 | 10 | 10 |  | 3 |  | 90 |
| Hospital visits | 10 | 31 | 15 | 9 | 18 | 2 | 1 | 2 | 88 |
| Chronic conditions | 2 | 12 | 8 | 1 | 4 |  | 2 |  | 29 |
| GP and community services | 7 | 7 | 5 | 2 | 2 | 1 | 2 | 1 | 27 |
| Personal care | 3 | 5 | 6 | 7 | 2 | 1 |  | 1 | 25 |
| Sensory issues | 1 | 8 | 5 | 2 | 2 | 3 | 3 |  | 24 |
| Oral health | 2 | 2 | 8 |  | 2 |  | 1 |  | 15 |
| Infection control | 1 | 2 | 3 | 1 | 2 |  |  |  | 9 |
| Covid19 | 2 | 1 | 2 |  | 1 |  | 1 |  | 7 |
| Sleep |  | 3 | 1 | 1 | 1 |  |  |  | 6 |
| Positioning |  |  |  | 2 | 2 |  | 1 | 1 | 6 |
| Frailty mentioned |  |  | 1 | 1 | 2 | 1 |  |  | 5 |
| **Quality of care & facilities** | 18 | 75 | 89 | 49 | 35 | 4 | 16 | 8 | 294 |
| Eating & drinking | 6 | 59 | 56 | 13 | 11 |  | 2 | 3 | 150 |
| Care safety & regulation | 8 | 7 | 14 | 10 | 13 |  | 7 | 5 | 64 |
| Care plans / documentation | 5 | 2 | 10 | 1 | 8 | 1 | 6 | 4 | 37 |
| Building design | 3 | 10 | 10 | 12 | 1 |  |  |  | 36 |
| Homeliness |  | 9 | 2 | 5 | 1 | 1 | 2 | 2 | 22 |
| Transport mentioned | 2 | 8 | 6 | 5 | 1 |  |  |  | 22 |
| "Person-centred care" mentioned | 1 | 1 | 10 | 1 | 1 |  | 3 | 1 | 18 |
| "Dignity" mentioned | 3 | 3 | 3 | 6 | 1 |  |  |  | 16 |
| Alternatives to care homes | 2 | 1 |  | 2 | 1 | 1 | 1 | 1 | 9 |
| Technology | 7 | 12 | 20 | 7 | 5 | 3 | 10 |  | 64 |
| AI mentioned | 1 |  | 3 |  | 1 | 1 |  |  | 6 |
| **Family involvement** | 11 | 20 | 22 | 29 | 10 | 1 | 12 | 3 | 108 |
| No Next of Kin | 1 | 4 | 1 | 3 |  |  | 1 |  | 10 |
| **Admissions/transitions** | 8 | 35 | 19 | 18 | 14 | 1 | 7 | 4 | 106 |
| **Funding & Finance** | 14 | 36 | 35 | 26 | 5 | 4 | 8 |  | 128 |
| Demographics (inc cultural diversity) | 1 | 22 | 22 | 5 | 5 |  | 4 |  | 59 |
| Request for information/learning | 5 | 17 | 14 | 12 | 5 |  | 2 |  | 55 |
| Public & media perception of CHs | 10 | 12 | 8 | 3 | 4 |  | 4 | 2 | 43 |
| Political decisions | 5 | 2 | 3 | 6 | 3 | 1 | 4 |  | 24 |
| Research methodology | 1 | 6 |  | 1 | 3 |  | 5 |  | 16 |

**Bold** denotes major themes, all others classed as subthemes

##### A11a Summary of less commonly reported themes

| **Theme** | **Examples** | **Example Quote** |
| --- | --- | --- |
| **End of life^1^** | Staff seek better training for residents who are dying and post-death care. Advance care planning ReSPECT forms were seen as essential but often poorly timed or explained. Avoiding inappropriate hospital admissions at the end of life was noted. Residents and staff expressed the need for grief support following loss of partners, homes, or fellow residents. | “End of life care - found to be hit and miss with staff having their own opinion's and bringing their own beliefs on palliative care. Not 'shipping' residents to hospital when end of life care is needed.” (Relative) |
| **Dementia^1^** | Priorities included improved family support, clearer understanding of different dementia types, and the relationship between dementia and depression. Staff wanted evaluation of dementia-awareness training and more non-pharmacological approaches (e.g. music, intergenerational activities). Interest was shown in links with neurodiversity and treatments such as CBD products and Melatonin. | “With the increased awareness of neuro diversity, I would like research to look at the link between this and dementia. like how many people have been through care services with undiagnosed neuro diversity but have been treat for other issues i.e dementia” (Staff) |
| **Funding & Finance^2^** | A major concern related to funding depletion and what this means for residents and their care. Residents and relatives emphasised the need for “value for money”, fair fees and improved processes in terms of accessing funding. Policymakers wanted to understand how funding models (self-funding vs local authority; regional variation) affect outcomes. | “Finances the most important concern - what happens when our money runs out?” (Resident)  “Correlation between care home fees and the quality of life of care home residents and staff.” (Owner/manager) |
| **Eating & hydration^3^** | Residents’ concerns included taste, appearance, and variety of modified consistency foods and the need for greater choice in menus (vegetarian, culturally diverse, nutritious). From health professionals, there was a call for better staff training on dietary needs, particularly related to IDDSI levels and swallow difficulties. Access to Speech and Language Therapy for swallow assessments seen as insufficient. | “Eating together. In most care homes, apart from sharing a bit of birthday cake, staff rarely sit down and eat together with residents. The potential benefits are huge, but this simple expression of common humanity is missed, and we are all poorer for it.” (Health care worker) |
| **Transition**  **into care2** | Key topics were better information and emotional support during the move into care, maintaining identity and privacy, and supporting changing family roles. Some wanted research on alternative models that help people stay at home, and on the purpose and review of respite/step-down placements. | “Joining care home is a stressful time - lots of unknowns. It would be very beneficial if homes could give new residents a booklet with all the information we need... Having it all down on paper would be helpful.” (Resident) |
| **Family involvement2** | Respondents raised challenges around families who are absent, overly reliant on the home, or unsure how involved to be. Strong calls were made for clearer communication and involvement in decisions, especially for residents with dementia or reduced capacity. Digital communication tools and dedicated liaison roles were suggested. | “Too much focus on paperwork, either with written or IT systems - older people and families never see the records” (Social services)  “Understanding finances and the guilt associated with moving a loved one into a care home” (Relative) |
| **Technology2** | Stakeholders recommended evaluating the impact of technology on care quality and efficiency. Suggested areas included falls-prevention tools, reminiscence technologies, and AI for administrative tasks. Barriers include staff digital skills, training needs and uneven uptake across homes. | “For families who live far away, the care home could offer a digital portal and video call stations, recorded video messages and a private family social media platform where updates, milestones and daily moments are shared” (Staff) |
| **Demographics2** | Shifting demographics included younger residents with modern food preferences and greater digital literacy. Cultural diversity needs included pronoun visibility, and language support. Some concerns were raised about inappropriate placements, mixing resident groups and managing conflict in shared spaces. | “Managing sexually inappropriate and disinhibited behaviours in residents with dementia.” (Health care worker)  “Should younger residents (in their 50s) be in units where people are 80+. I feel the dynamics don't work” (Resident) |

1 **Subtheme of Health & Medical**
2 **Major Theme**
3 **Subtheme of Quality of Care**

##### A11b Viable research questions compared to volume of discussion

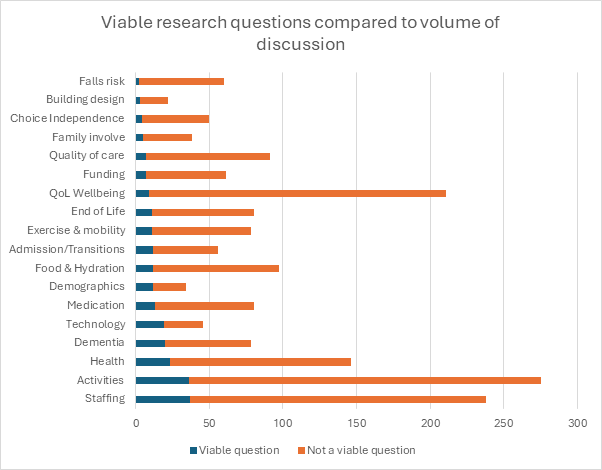

##### A11c Suggested themes & total number of mentions

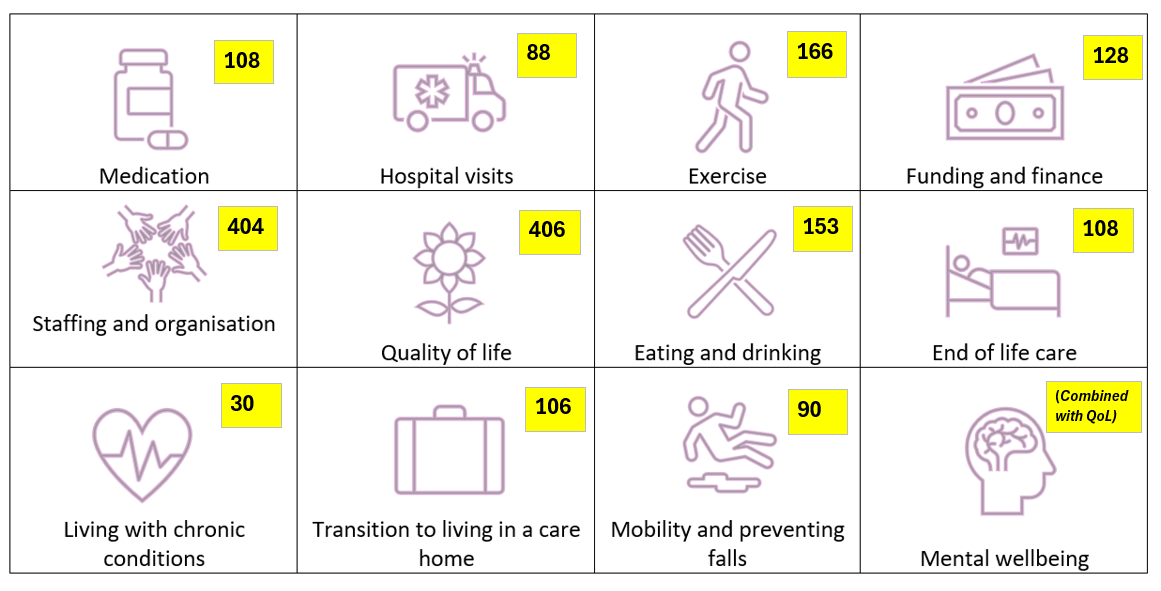

*The "Priorities Poster" was a resource included in the activity packs to help explore research priorities with residents. It was also used in the online survey. It presented a list of suggested topics for researchers to focus on, designed to stimulate discussion and encourage participants to think about what could be improved. Residents could indicate their preferences by saying, pointing to, or using stickers/stamps on the areas most important to them. The poster, along with detailed answers on suggestion cards, was returned by fourteen care homes. Note that our most commonly reported priority – Engagement with activities (n=443) was not explicitly listed.*

##### A12 Priority ranking heat map–Stakeholders & previous studies

|  | **General theme** | **PREACH Residents (n=381)** | **PREACH Staff (n=346)** | **PREACH Family (n=139)** | **PREACH Health care (n=70)** | **PREACH Manager/owners (n=62)** |  | **Search-U (2025) (Res, Rel & Staff)** | **ENRICH Scotland 2014,2022,2024 (staff)** | **Rayner (2019) (staff)** | **Gordon (2018) (academics/health p)** | **Shepherd (2017) (staff)** |
| --- | --- | --- | --- | --- | --- | --- | --- | --- | --- | --- | --- | --- |
|  | **Engagement with activities** | **1st** | **3rd** | **3rd** | **3rd** | **4th** |  | **1st** | **3rd** | **1st** |  |  |
|  | **Staffing** | **4th** | **2nd** | **2nd** | **2nd** | **1st** |  |  | **2nd** |  |  | **3rd** |
|  | **Quality of life and wellbeing** | **2nd** | **1st** | **1st** |  | **3rd** |  |  |  |  |  |  |
|  | **Health or medical** | **3rd** | **4th** |  | **1st** | **2nd** |  | **2nd** | ***1st** |  |  |  |
|  | **Quality of care & facilities** |  |  | **4th** | **4th** |  |  |  |  |  | **1=** |  |
|  | **Eating & drinking** |  |  |  |  |  |  |  |  |  | **1=** |  |
|  | **Dementia** |  |  |  |  |  |  |  | ***1st** | **2nd** |  |  |
|  | **Funding & Finance** |  |  |  |  |  |  |  |  | **3rd** |  |  |
|  | **Admissions/transitions** |  |  |  |  |  |  |  |  |  |  |  |
|  | **End of life care** |  |  |  |  |  |  | **3rd** |  |  | **1=** |  |
|  | **Hospital visits** |  |  |  |  |  |  |  |  |  | **1=** |  |
|  | ***Person-centred care*** |  |  |  |  |  |  |  |  |  |  | **1st** |
|  | ***Dignity*** |  |  |  |  |  |  |  |  |  |  | **2nd** |

##### A13 Potential Interventional / RCT Ideas

| **Stakeholder group** | **Priority / Idea "My idea for care home research is…"** |
| --- | --- |
| Care home owner or manager | Technology: Baseline metrics of where we are not using technology vs difference after implementing technology |
| Care home owner or manager | Research into how individualised exercise plans improve mobility, balance and fall prevention in care home residents. Compare the effectiveness of social group exercise versus one-on-one physiotherapy sessions.  Research into whether structured physical activity helps to slow cognitive decline in residents with dementia. Identify factors that prevent care home residents from engaging in physical activities and potential solutions. |
| Care home owner or manager | How non-pharmacological interventions affect agitation in residents with dementia |
| Care home resident | More doll therapy and for carers to do more activities on units when activities cant be done by activity workers |
| Care home resident | Animals impact on wellbeing - could therapy animals visit & impact of this be measured |
| Care home resident | Mobility and preventing falls - how to protect vulnerable residents - can other residents help - be trained / educated? |
| Care home staff member | The effectiveness of dementia awareness training on staff attitudes and behaviors towards residents . |
| Care home staff member | Use of SAD lamps Water therapy - floation, physio in a pool Exercise classes - who does what (eg seated, exercise bikes) and what is the benefit? |
| Care home staff member | Benefits of exercise and strengthening activities to aid movement, walking, gross motor (dressing independently), measuring improvements |
| Care home staff member | Look at the healing of wounds - how effective is adding protein to someone with a wound? |
| Care home staff member | How effective is adcal for someone with osteoporosis? |
| Care home staff member | Is it true that supplements like creatine may help factors of dementia? |
| Care home staff member | Nutrition, eating & drinking - hydration jellies - are they evidenced? |
| Family or friend of care home resident | Can exercise and physio be made a regular, extended, compulsory and publicly evidenced activity as it can seem residents are all to easily left to live in a chair or bed and given up on? |
| Family or friend of care home resident | Explore the benefits and challenges of intergenerational programs within care home settings |
| Family or friend of care home resident | Assess the role of sensory gardens for residents with cognitive impairments |
| Health care worker | Eating and drinking abilities, access to speech and language therapy (SALT) for swallow assessments, training from SALT to identify swallow difficulties and how to support this client group with difficulties, waiting times for SALT assessments in community. There is no research outlining needs for SALT input in community to help us emphasises staffing needs to CCGs. Staffing gets emphasised for hospital teams not community. |
| Health care worker | Physiotherapy. Care home residents are statistically higher risk of falls and there is a clear evidence based link between falls reduction and physiotherapy. A regular assessment from a qualified physiotherapist or physiotherapist trained assistant should be part of residential care. |
| Health care worker | Use of Melatonin in care home settings, particularly dementia. We have seen extraordinary benefits from Melatonin for PWD in care homes - not everyone responds but those who do derive a significant benefit. Unfortunately this is not recommended by NICE nor always readily accessible through primary care if at all and robust research into this would potentially support significant improvements in the sleep, physical and mental wellbeing of residents as well as reduce challenging behaviour and carer burden - while not increasing the risk of falls, delirium and anticholinergic burden that other hypnotic medications or medications for challenging behaviour can. |
| Health care worker | Maintaining continence/preventing UTIs - links to preventing infections that can lead to sepsis, general QoL and well-being, maintaining dignity and personal hygiene standards |
| Health care worker | Chronic Obstructive Pulmonary Disease. Nice guidance and Gold Standard guidance suggests quick and easy access to steriods and/or anti-biotics for people living with COPD. This does not happen for those in care homes. XXX Locally staff have been trained with a protocol in place to give guidance as to the start of an infection for a resident with COPD. Hospital admission reduced, as did the need for chest infection. |
| Policy & other | Introducing care robots to be the second care worker or nurse carrying out personal care, manual handling, mobilisation, recording of care and medication dispensing and auditing. They would also be able to report abuse or poor practice and potentially facilitate communication with friends and families |
| Researcher | Examine intergenerational programs and their benefits for residents with dementia |
| Researcher | Use of legal CBD products in reducing stress/distress other symptoms for residents living with Dementia |
| Researcher | Benefits of structured routine exercise in care homes (e.g. daily morning walk) on quality of life, mood, anxiety & health outcomes (RCT design) |

##### A14 Potential for Secondary data analysis

| **Stakeholder group** | **Priority / Idea "My idea for care home research is…"** |
| --- | --- |
| Care home owner or manager | Research into the impact of reducing unnecessary medications on residents health and quality of life. Research into how targeted training improves care staffs confidence and accuracy in administering medications. Explore how regular pharmacist led reviews influence health outcomes and hospital admissions. |
| Care home owner or manager | Correlation between care home fees and the quality of life of care home residents and staff. |
| Care home owner or manager | What is the national average of medication errors within residential settings, and what is the impact to overall care delivery that medications errors have? |
| Care home owner or manager | Correlation between number of hours of GP visit to the care home and health outcomes to the care home residents (e.g. hospital admissions, place of death, infections...) |
| Care home owner or manager | The correlation between the use of anti psychotic medications and providing therapeutic activities in the elderly. |
| Care home resident | Compare care homes so other homes know what is being done and can share best practice |
| Care home resident | percentage of care workers who are encouraged to do a higher level at college or do a nursing degree |
| Care home staff member | How medication impact, communication and Mobility. |
| Care home staff member | What is the most common age range people die? |
| Care home staff member | We try to book transport to take residents to hospital but it gets cancelled or they have to wait hours. It feels to us like often because residents are in a 'safe place' they get treat as second class citizens. When it come to ambulances, hospital transport, GP appointments, everything. I would like to see if there is evidence this is the case and how do we improve it |
| Care home staff member | Will we ever have an 'agreed safe staffing level for care homes'? There is no specific and clear guidance and the staffing ratios are typically set by those who do not work 'on the floor' and who are more focused on 'staffing costs', dependencies are carried out but quite often these are not reflective and do not take in to account all true aspects of daily care delivery with some being very outdated yet still in use. Who checks that the staffing levels laid down by Manager are indeed safe and appropriate? |
| Care home staff member | What is the average time after someone is diagnosed with dementia last upon? |
| Care home staff member | Staff to resident ratios, staff survey of job satisfaction, resident needs for standard and complex payments, fees comparison over the regions  Support from local councils, admission reasons to hospital, falls, dehydration, pressure sores |
| Care home staff member | more in-depth research into coms(?), meds, quality of life and transitioning |
| Family or friend of care home resident | What is the degree of medication? Is it all necessary as many meds have issues attached. A useful area for research might be to find attitudes among care homes, GPs, and patients to reduction of medication? With further work to find what results when reduction is done? I would anticipate cost savings in drug provision for the NHS and the home (in terms of labour), but most of all, improvement in QoL for residents. |
| Family or friend of care home resident | Individuals in care settings who have capacity, and require medication should have the right to choose to self-administer prescribed medication where they are able to do so.  How can this be effectively and safely managed in order to meet the regulations around medication storage and administration? |
| Health care worker | im a GP I want focussed research on this age group to be practical around medication is it worth continuing antihypertensives? what about statins? DOACs and falls risk in the very frail |
| Health care worker | How can we use existing resources to minimise admissions to hospital for care home residents |
| Health care worker | I would like to compare outcomes of patients who attended hospital and those who didnt lots of anecdotal evidence that outcomes are better when you stay away from secondary care. is this true? |
| Health care worker | Many care home residents may be lost to follow up from specialist clinics because they are no longer able to attend e.g. Parkinsons patients. How many people with PD are living in care homes and when were they last reviewed by an PD nurse/doctor? Are their end of life needs being met? |
| Health care worker | In terms of national policy direction for the development of Urgent Community Response Teams, Virtual Wards and Enhanced Health for Care Homes - there needs to be a research evaluation approach to understanding delivery models and interface with care homes e.g. I know of one GP who spends 2 hours in a care home and another who spends 8 - what are the outcomes - variance in practice might be fine but if the priority for the policy is e.g. to reduce unscheduled care use through improving proactive care via 'home rounds' then how do related metrics look when comparing variance in delivery - might be fine - might not be.... |
| Policy & other | The inequalities of how care is funded e.g. fully funded CHC, private care and social services with top up contribution. |
| Researcher | Optimisation and reduction of medications for older adults in treating health conditions (eg multiple medications for one condition such as high blood pressure) as residents take a large number of medications which can often lead to refusals especially with cognitive impairments |
| Researcher | Understanding the heterogeneity within the care home population and how clusters of characteristics affect outcomes. More than looking at the cluster of diseases, for example, how entry point ( hospital, home, residential care) affects outcomes, self-funding vs LA support, functional ability etc. |
